# Characterization of mitochondrial DNA quantity and quality in the human aged and Alzheimer’s disease brain

**DOI:** 10.1101/2021.05.20.21257456

**Authors:** Hans-Ulrich Klein, Caroline Trumpff, Hyun-Sik Yang, Annie J. Lee, Martin Picard, David A. Bennett, Philip L. De Jager

## Abstract

Mitochondrial dysfunction is a feature of neurodegenerative diseases, including Alzheimer’s disease (AD). Using whole-genome sequencing, we assessed mitochondrial DNA (mtDNA) heteroplasmy levels and mtDNA copy number (mtDNAcn) in 1,361 human brain samples of five brain regions from three studies. Multivariable analysis of ten common brain pathologies identified tau pathology in the dorsolateral prefrontal cortex and TDP-43 pathology in the posterior cingulate cortex as primary drivers of reduced mtDNAcn in the aged human brain. Amyloid-β pathology, age, and sex were not associated with mtDNAcn. Further, there is evidence for a direct effect of mitochondrial health on cognition. In contrast, while mtDNA heteroplasmy levels increase by about 1.5% per year of life in the cortical regions, we found little evidence for an association with brain pathologies or cognitive functioning. Thus, our data indicates that mtDNA heteroplasmy burden is unlikely to be involved in the pathogenesis of late-onset neurodegenerative diseases.

## Introduction

Mitochondria are complex multi-functional organelles involved in various pathways including fatty acid and cholesterol synthesis, apoptosis, calcium signaling, and adenosine triphosphate generation (Nunnari and Suomalainen, 2012; Spinelli and Haigis, 2018). Dysfunctional mitochondria have been described in aging (Sun et al., 2016) and in many neurodegenerative diseases such as Alzheimer’s disease (AD) and amyotrophic lateral sclerosis (ALS) (Area-Gomez et al., 2019). Mitochondria harbor their own circular genome of 16,569 base pairs encoding 13 proteins of the respiratory chain. Mitochondrial DNA (mtDNA) can be replicated independent of the cell cycle. Since mtDNA expression is required for respiratory activity, the mtDNA copy number (mtDNAcn) within a cell is regulated to meet the cell’s metabolic needs, resulting in a wide range of mtDNAcn in different tissues and conditions (D’Erchia et al., 2015; Filograna et al., 2020). Like nuclear DNA, mtDNA can also carry mutations, which either affect all copies of the mtDNA in a cell (termed homoplasmy) or only a fraction of the mtDNA molecules (termed heteroplasmy). Heteroplasmic variants are assumed to be somatically generated or inherited as low level variants, and they can clonally expand over an individual’s life-time (Stewart and Chinnery, 2021).

The mtDNAcn has become a popular potential marker of mitochondrial health in translational studies, because mtDNAcn can be measured in stored biospecimens at large scale using qPCR or DNA sequencing techniques. In AD, several studies have investigated mtDNAcn in tissue homogenates from different brain regions and found either a lower mtDNAcn in AD or no significant changes. One of the strongest reductions (50%) was reported by an early study of the frontal cortex (Coskun et al., 2004). Smaller effect sizes or non-significant changes were reported for the hippocampus, cerebellar cortex, and cerebellum, indicating the possibility of brain region-specific effects (Rice et al., 2014; Rodriguez-Santiago et al., 2001; Wei et al., 2017). Similar results were reported for other neurodegenerative diseases (Filograna et al., 2020). Interestingly, although mtDNAcn derived from whole blood is often confounded by variation in cell type composition between individuals, a recent study found an association with AD suggesting that the mtDNAcn in whole blood could potentially reflect metabolic health across tissues (Yang et al., 2021). While the mtDNAcn overall seems to be reduced in brain regions affected by neurodegenerative diseases, mixed results have been reported for the effect of aging on mtDNAcn. For example, two studies found no evidence for age-related changes of mtDNAcn in three brain regions, skeletal muscle, and heart muscle (Frahm et al., 2005; Miller et al., 2003), whereas a more recent study reported a decrease in skeletal muscle and an increase in liver tissue with age (Wachsmuth et al., 2016).

A higher burden of mtDNA heteroplasmy has been observed in tissues from aged individuals (Larsson, 2010). Increased levels of mtDNA heteroplasmy were also described in brains from AD and Parkinson’s disease (PD) patients (Bender et al., 2006; Coskun et al., 2004), which lead to the hypothesis that pathogenic mtDNA mutations, when they exceed certain thresholds, could contribute to the mitochondrial dysfunction observed in late-onset neurodegenerative diseases. However, other studies found no evidence for an association between mtDNA heteroplasmy levels and AD or PD (Wei et al., 2017).

In this study, we profiled mtDNAcn and mtDNA heteroplasmy levels in n=762 brain samples from the Religious Orders Study and the Rush Memory and Aging Project (ROSMAP) (Bennett et al., 2018) and implemented substantial improvements compared to previous studies: (i) The detailed pathologic characterization of ROSMAP samples facilitated the disentanglement of the effects of different brain pathologies and aging on mtDNA. Mixed pathologies are common in aged individuals including AD patients (Schneider et al., 2007) and unaccounted pathologies may have contributed to some of the mixed results in the literature. (ii) Standardized cognitive tests conducted proximate to death were employed to assess the association between cognitive functioning and mtDNAcn adjusted for pathologies. (iii) Using RNA-seq-derived estimations of cell type proportions, we accounted for neuronal loss as a major confounder of mtDNAcn analyses in AD brains. (iv) We profiled five different brain regions to assess brain-regional differences (three regions in ROSMAP, and two additional regions in two independent datasets with a total of n=599 additional samples). (v) Finally, we calculated a proteomic score representing mitochondrial content to investigate whether changes in mtDNAcn reflect changes of mitochondrial mass or whether they are specific to mtDNA maintenance.

## Results

### mtDNAcn is reduced in the AD cortex

Whole genome sequencing (WGS) data were generated from n=762 post mortem brains from ROSMAP, two harmonized cohort studies of aging and dementia (Bennett et al., 2018). Brain specimens were obtained from the dorsolateral prefrontal cortex (DLPFC) (n=454), the posterior cingulate cortex (PCC) (n=66), and the cerebellum (CB) (n=242) from different individuals. Table 1 summarizes pathologic and demographic information of the samples. The number of aligned sequence reads per sample ranged from 643.5×10^6^ to 1,475.0×10^6^ reads (median of 899.1×10^6^ reads) (Supplementary Excel File 1). The mtDNAcn was estimated from the WGS data by dividing the median coverage of the MT chromosome by the median coverage of the autosomal chromosomes and multiplying the ratio by two (Ding et al., 2015; Longchamps et al., 2020).

**Table 1.** Characteristics of the ROSMAP cohort.

|  | DLPFC (n=454) | PCC (n=66) | CB (n=242) |
| --- | --- | --- | --- |
| Sex (male) | 158 (34.8%) | 21 (31.8%) | 71 (29.3%) |
| Age (years) | 89.3 (6.6) | 89.1 (5.6) | 88 (6.7) |
| Pathologic AD | 308 (67.8%) | 45 (68.2%) | 141 (58.3%) |
| Amyloid (% area affected) | 4.6 (4.4) | 3.9 (3.8) | 4 (4.3) |
| Tau (% area affected) | 7.4 (8.1) | 6.8 (6.8) | 6.2 (7.9) |
| TDP-43 (none, amygdala, limbic, neocortical) | 205, 76, 97, 44<br>(48.6%, 18.0%, 23.0%, 10.4%) | 23, 16, 12, 12<br>(36.5%, 25.4%, 19.0%, 19.0%) | 96, 45, 38, 33<br>(45.3%, 21.2%, 17.9%, 15.6%) |
| Lewy bodies (none, nigral, limbic, neocortical) | 337, 7, 32, 60<br>(77.3%, 1.6%, 7.3%, 13.8%) | 48, 2, 2, 9<br>(78.7%, 3.3%, 3.3%, 14.8%) | 184, 6, 21, 25<br>(78.0%, 2.5%, 8.9%, 10.6%) |
| Cerebral amyloid angiopathy (none, mild, moderate, severe) | 92, 191, 110, 52<br>(20.7%, 42.9%, 24.7%, 11.7%) | 13, 25, 16, 11<br>(20.0%, 38.5%, 24.6%, 16.9%) | 58, 101, 51, 24<br>(24.8%, 43.2%, 21.8%, 10.3%) |
| Cerebral atherosclerosis (none, mild, moderate, severe) | 84, 212, 123, 33<br>(18.6%, 46.9%, 27.2%, 7.3%) | 11, 25, 19, 11<br>(16.7%, 37.9%, 28.8%, 16.7%) | 39, 114, 72, 15<br>(16.2%, 47.5%, 30.0%, 6.2%) |
| Arteriolosclerosis (none, mild, moderate, severe) | 136, 157, 118, 42<br>(30.0%, 34.7%, 26.0%, 9.3%) | 9, 26, 22, 9<br>(13.6%, 39.4%, 33.3%, 13.6%) | 78, 79, 65, 17<br>(32.6%, 33.1%, 27.2%, 7.1%) |
| Gross chronic infarcts (one or more) | 171 (37.7%) | 22 (33.3%) | 83 (34.3%) |
| Chronic microinfarcts (one or more) | 132 (29.1%) | 18 (27.3%) | 70 (28.9%) |
| Hippocampal sclerosis (present) | 39 (8.7%) | 6 (9.2%) | 22 (9.2%) |
| Post mortem interval (hours) | 8.4 (5.7) | 6.6 (4.7) | 8.3 (6.7) |

Column headers denote the brain regions where specimens for WGS were sampled. Reported pathology burdens were obtained by considering multiple regions (see methods) and are not specific to the brain region selected for WGS. Mean and standard deviation are shown for continuous variables. Absolute frequency and percentage are shown for categorical variables.

We first studied whether the mtDNAcn in the three selected brain regions was altered in individuals with pathologic AD diagnosis compared to those without AD (Fig. 1A-D). Two different DNA extraction kits were used for the DLPFC samples. Since the DNA extraction method can influence the mtDNAcn estimates (Guo et al., 2009; Nacheva et al., 2017), the DLPFC samples were split by kit and analyzed separately. In line with previous reports (Coskun et al., 2004; Wei et al., 2017), we found a lower mtDNAcn in AD in the DLPFC (p=0.0029 and p=0.040, Wilcoxon rank-sum tests) and also in the PCC (p=0.055, Wilcoxon rank-sum test). Changes in the PCC were of similar magnitude but failed to reach significance probably due to the smaller sample size. Changes in the CB were not significant.

**Figure 1.**
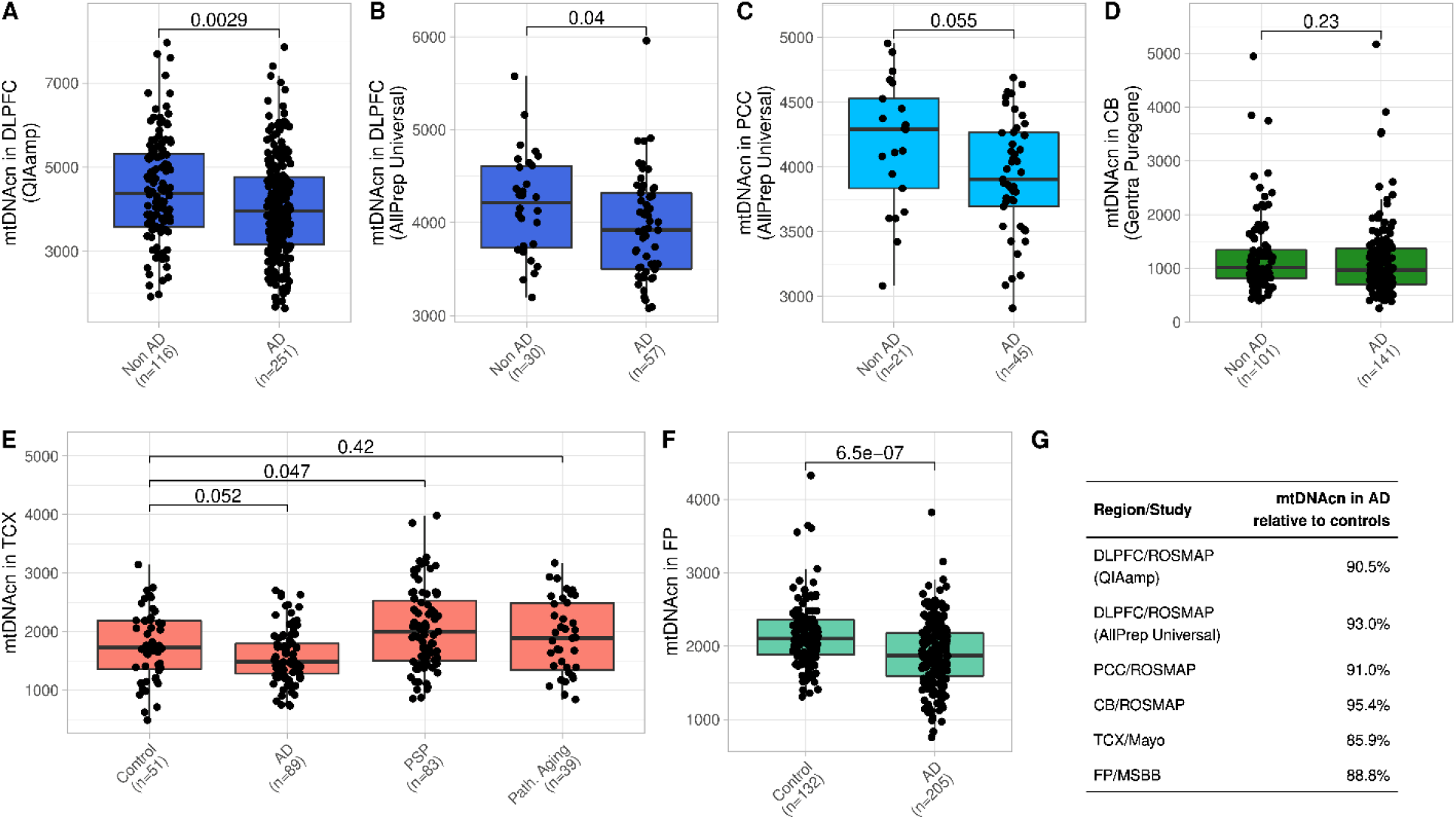
The mtDNAcn is reduced in cortical brain regions in AD. (A-D) Boxplots comparing the mtDNAcn in the DLPFC (A and B), in the PCC (C), and in the CB (D) from individuals of the ROSMAP cohort with and without pathologic AD diagnosis. The used DNA extraction kit is denoted in brackets on the y axis. Wilcoxon rank-sum test was applied to calculate p values. (E) Boxplot shows the mtDNAcn in the TCX of controls, AD cases, PSP cases, and cases of pathologic aging from the Mayo study. Wilcoxon rank-sum test was applied to calculate p values. (F) Boxplot shows the mtDNAcn in the FP of controls and AD cases from the MSBB study. Wilcoxon rank-sum test was applied to calculate p values. (G) The estimated relative mtDNAcn observed in AD compared to controls is shown for each brain region and study.

Next, we analyzed the mtDNAcn in two additional brain regions using public WGS data from the Mayo study and the Mount Sinai Brain Bank (MSBB) study (Tables S1,S2). Both regions, the temporal cortex (TCX) in the Mayo data (p=0.052, Wilcoxon test) and the frontal pole (FP) in the MSBB data (p=6.5×10^−7^, Wilcoxon test), demonstrated a lower mtDNAcn in AD samples compared to control samples (Fig. 1E,F). The reduction of mtDNAcn in AD ranged from 7.0% to 14.2% across the cortical regions from the three studies (Fig. 1G). The Mayo study includes samples diagnosed with pathologic aging, which is characterized by amyloid-β loads at similar levels as in AD and an absence of tau pathology. Whether pathologic aging is an early stage of AD or whether these individuals have protective factors that prevent the development of tau pathology is unknown (Murray and Dickson, 2014). We did not observe altered mtDNAcn levels in the TCX of individuals with pathologic aging (Fig. 1E). Additionally, the Mayo study includes samples with progressive supranuclear palsy (PSP). PSP is a tauopathy primarily characterized by tau inclusions in the brain stem and subcortical neurons. Interestingly, we observed a moderately higher mtDNAcn in the TCX of the PSP samples compared to the controls (p=0.047, Wilcoxon rank-sum test) (Fig. 1E).

For the subsequent analyses, we log-transformed the raw mtDNAcn and calculated z-scores for each of the six datasets shown in Fig. 1A-F. Then, we merged the two ROSMAP DLPFC datasets generated by different DNA extraction kits and verified that the normalized mtDNAcn approximately follows a standard normal distribution (Fig. S1). We note that the raw mtDNAcn before normalization should also be considered as a relative rather than an absolute measurement since the estimations are likely affected by experimental factors, which impedes a comparison across different brain regions or studies when different protocols and reagents were used.

### Lower mtDNAcn is primarily related to tau in the DLPFC and to TDP-43 in the PCC

AD pathology is the most common brain pathology in the aged brain but is often accompanied by comorbid brain pathologies. To determine which pathological feature is driving the association with mtDNAcn, we first analyzed each of the 12 pathologic variables shown in Fig. 2A as well as cognition and cognitive decline separately adjusted only for age and sex. The two cognitive variables were additionally adjusted for education. Further, we also analyzed the association with age and sex adjusted for AD pathology (Fig 2A). Detailed results of these analyses are given in Table S3 and can be summarized by four main findings: First, in the DLPFC, mtDNAcn is mainly associated with tau (p=2.9×10^−6^, t test) rather than amyloid-β pathology (p=0.011, t test) and is also associated with cognition proximate to death (p=7.9×10^−10^, t test) and cognitive decline (p=1.8×10^−9^, t test). Second, in the PCC, we also found an association with cognition (p=1.7×10^−5^, t test) and cognitive decline (p=5.5×10^−4^, t test), but the association with tau was not significant probably due to the smaller sample size. Interestingly, we found an association with TDP-43 pathology (p=2.1×10^−3^, F test) in the PCC, which explained 23% of the variance in mtDNAcn. Third, in the CB, neither pathologies nor cognitive measures were associated with mtDNAcn. Fourth, neither age nor sex were associated with mtDNAcn in any of the three regions when adjusting for AD pathology. All significant pathologies demonstrated an inverse correlation with mtDNAcn, and a lower mtDNAcn was associated with lower cognitive performance and a steeper rate of cognitive decline, consistent with the notion that a high mtDNAcn is a feature of healthy mitochondrial function in the aged brain.

**Figure 2.**
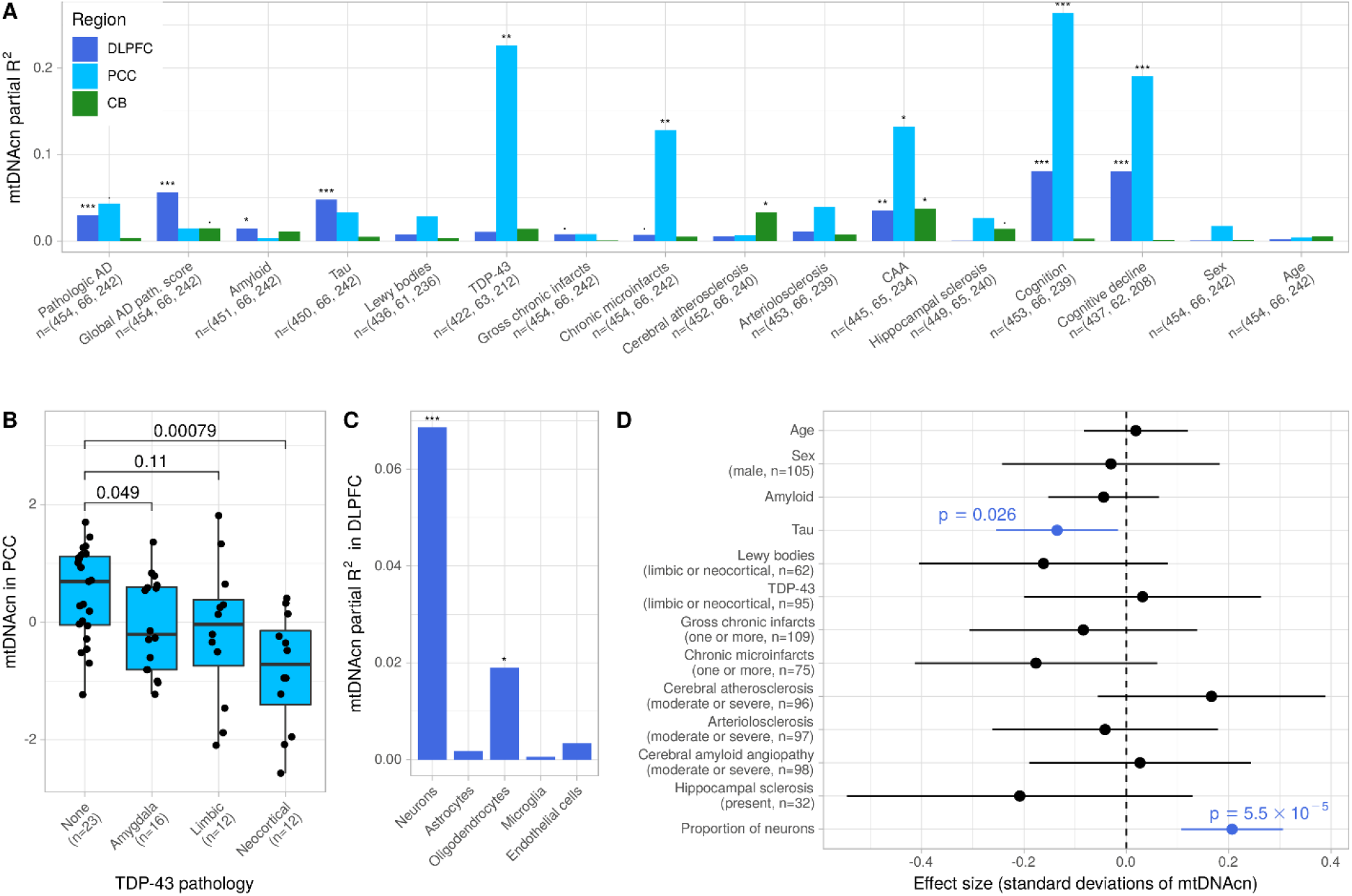
Changes of the mtDNAcn are primarily associated with tau in the DLPFC and TDP-43 in the PCC. (A) The bars indicate the mtDNAcn’s variance explained (partial R^2^) by different pathologies, cognitive measures, and demographics in the ROSMAP cohort. Each Variable was analyzed separately in a regression model with mtDNAcn as outcome adjusted for sex and age. The cognitive variables were additionally adjusted for education. The results for sex and age were obtained from a model adjusted for global AD pathology. Colors indicate brain regions. Asterisks indicate significance levels obtained by F-tests (*** for p ≤ 0.001, ** for p ≤ 0.01, * for p ≤ 0.05, and ? for p ≤ 0.1). The sample size available for each pathology and brain region is denoted on the y axis. (B) Boxplot shows the mtDNAcn in the PCC for different stages of TDP-43 pathology. Wilcoxon rank-sum test was used to calculate p values. (C) The bars indicate the mtDNAcn’s variance explained (partial R^2^) by cell type proportions estimated from RNA-seq data in a subset of n=327 DLPFC samples. Each cell type proportion was analyzed separately in a regression model with mtDNAcn as outcome adjusted for sex and age. Significance levels were obtained and indicated by asterisks as in (A). (D) Forest plot shows the result from a multivariable regression model with mtDNAcn in the DLPFC as outcome and the pathologic and demographic variables denoted on the y axis as explanatory variables. Estimated coefficients are shown as dots and the line segments represent the respective 95% confidence intervals. Continuous variables were z-standardized. Categorical variables were dichotomized and the factor level and case numbers corresponding to the plotted coefficient are denoted in brackets under the variable name. t-test was applied to calculate p values. A total of n=288 cases with complete observations of all variables were used to fit the model.

Pathologic TDP-43 is known to localize inside mitochondria and has recently been described to trigger the release of mtDNA molecules from the mitochondrial matrix into the cytoplasm in cellular systems (Wang et al., 2013; Wang et al., 2016; Yu et al., 2020). In our study, limbic-predominant age-related TDP-43 encephalopathy neuropathological change (LATE-NC) was captured by phosphorylated TDP-43 immunohistochemistry, and staged according to the recent consensus working group report (stage 0, no TDP-43; stage 1, localized to amygdala; stage 2, extension to hippocampus or entorhinal cortex; stage 3, extension to the neocortex) (Nelson et al., 2019). In the PCC, we observed a gradual reduction of mtDNAcn across the stages with a distinctly lower mtDNAcn in stage 3 (Fig. 2B). In contrast, in the DLPFC we observed only a minor reduction in the latest stage (Fig S2A). Our findings are consistent with previous reports of prominent PCC hypometabolism in LATE-NC (Botha et al., 2018), and DLPFC being involved only in the most advanced stage of LATE-NC (Josephs et al., 2016; Nag et al., 2018). Thus, PCC neurodegeneration may be an early feature of LATE-NC, even though the current LATE-NC staging scheme does not consider the PCC.

Neurons have relatively high concentrations of mitochondria, and changes in the proportion of neurons during the course of AD can confound mtDNAcn measurements at the tissue level. We therefore estimated the proportions of neurons and of glial cell types from DLPFC RNA-seq data for a subset of n=327 samples with DLPFC mtDNAcn estimates. As expected, the neuronal proportion was significantly positively associated with mtDNAcn, accounting for 6.9% of the variance in mtDNAcn (p=1.7×10^−6^, t test) (Fig. 2C, Table S4). We also found an association with oligodendrocytes (p=0.013, t test), but in contrast to the neuronal proportion this association disappeared when we modeled the different cell types together, indicating that adjusting for neuronal proportions in downstream analyses is sufficient.

Since many of the pathologies are correlated with each other, we next modeled all pathologies, age, sex and neuronal proportion in a multivariable regression model to identify the primary driver of mtDNAcn changes in the DLPFC. As shown in Fig 2D, only tau pathology and neuronal proportion remained significantly associated with mtDNAcn when accounting for all pathologies. Thus, our data suggest that tau burden is directly associated with lower mtDNAcn in the DLPFC and that this relationship cannot be explained by neuronal loss alone. Similar results for tau were obtained from a model unadjusted for neuronal proportion (Fig. S2B).

### mtDNAcn affects cognitive functioning independent of brain pathologies

The univariate analysis (Fig. 2A) revealed that a lower mtDNAcn in the cortex is associated with a decline of cognitive function. We next sought to investigate whether this relation can be explained by the correlation of mtDNAcn with tau, or whether mtDNAcn and tau have effects on cognitive functioning independent of each other. We modeled cognitive function as outcome depending on age, sex, education, mtDNAcn, and ten neuropathologies including tau using our DLPFC data. In this model, mtDNAcn was significantly associated with cognitive function (p=2.4×10^−4^, t test) together with tau, Lewy bodies, gross chronic infarcts, hippocampal sclerosis, and education (Fig 3A). Tau was the most important predictor for cognitive function. When we added neuronal proportion to the model, the effect size of mtDNAcn was attenuated by 26% but remained significant (p=0.034, t test) despite the smaller number of samples that have both measurements neuronal proportion and mtDNAcn (Fig. 3B). These results indicate that the observed changes in the mtDNAcn in the DLPFC capture aspects of mitochondrial health beyond neuronal loss and affect cognitive functioning independent of pathologies.

**Figure 3.**
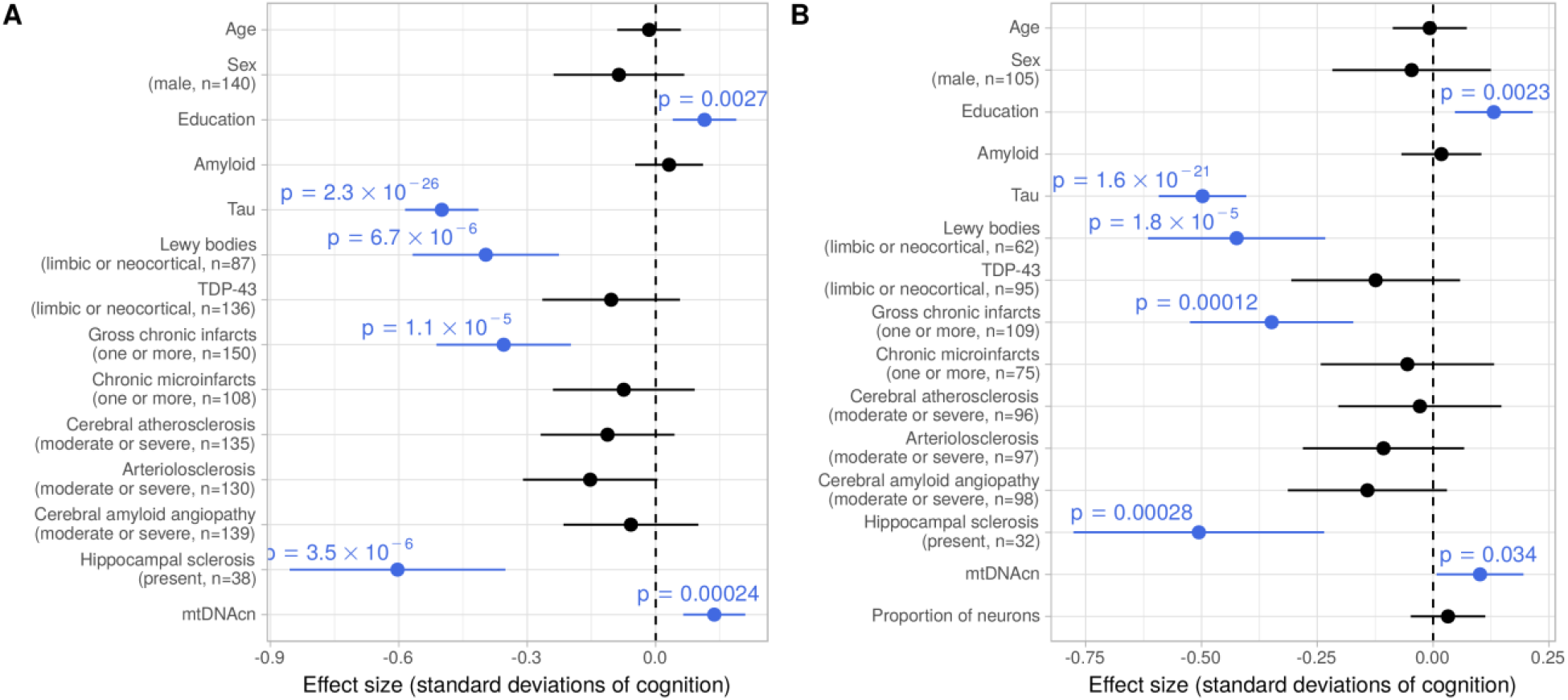
The mtDNAcn is associated with cognitive function independent of brain pathologies. (A) Forest plot shows the result from a multivariable regression model with global cognitive function as outcome and the pathologic variables, demographic variables and mtDNAcn as explanatory variables (y axis). Estimated coefficients are shown as dots and the line segments represent the respective 95% confidence intervals. Continuous variables were z-standardized. Categorical variables were dichotomized and the factor level and case numbers are denoted in brackets under the variable name. t-tests were applied to calculate p values. A total of n=393 cases with complete observations of all variables were used to fit the model. (B) Same plot as in (A) but with neuronal proportion as additional variable in the model reducing the number of samples with complete observations to n=287.

### Genetic determinants of the mtDNAcn

Two recent genome-wide association studies (GWAS) of blood samples from the UK Biobank identified 96 and 50 independent loci respectively that affected the mtDNAcn in whole blood (Hagg et al., 2020; Longchamps et al., 2021). Functional annotation of some loci pointed to blood-specific pathways such as platelet activation or megakaryocyte proliferation, an association consistent with the influence of platelets on whole blood mtDNAcn (Shim et al., 2020), whereas other loci were located at genes involved in mitochondrial pathways and therefore could regulate the mtDNAcn in brain tissues as well. Since our sample size is limited for a GWAS, we conducted a focused analysis of 81 lead SNPs identified in the prior study (Longchamps et al., 2021) and with a minor allele frequency ≥ 5% in our data. We separately analyzed the four brain regions with at least 100 samples (DLPFC, CB, TCX, and FP) and subsequently performed a random effects meta-analysis. A total of n=67 non-Caucasian individuals in the MSBB dataset (FP) were excluded resulting in n=1,228 brain samples. The association tests were adjusted for age, sex, population structure, and quantitative AD pathology score (DLPFC, CB) or post mortem diagnosis respectively (TCX, FP). After Bonferroni adjustment, only the top SNP rs11085147 from the original study was significantly associated with mtDNAcn in our brain data (p_BF_=0.0436). The direction of the effect was identical in all four brain regions and consistent with the originally reported direction in blood. Each additional dosage of the alternative allele increased the mtDNAcn by 0.23 standard deviations. The SNP rs11085147 is a missense variant of the gene Lon Peptidase 1, Mitochondrial (*LONP1*), which binds mitochondrial DNA and is involved in mtDNA replication and mitogenesis (Bota and Davies, 2016). rs11085147 has not been linked to AD risk in genome-wide association studies of AD. Detailed results for all 81 SNPs are given in Supplementary Excel File 2.

The strongest genetic risk factor for late-onset Alzheimer’s disease is the Apolipoprotein E (*APOE*) locus. Several studies using mouse models or human brain implicated the ε4 risk allele with mitochondrial dysfunction (Area-Gomez et al., 2020; Yin et al., 2020). We therefore analyzed the effect of the *APOE* ε4 allele on the mtDNAcn in our four datasets with ≥100 samples. The *APOE* ε4 allele was associated with lower mtDNAcn (p=8.0 × 10^−7^, random effects meta-analysis), but this association was attenuated considerably when adjusting for pathologies (p=0.014, random effects meta-analysis) (Table 2) suggesting that a large fraction but not the complete *APOE* e4 effect on mtDNAcn is mediated via AD pathology. Indeed, mediation analysis of the DLPFC samples (Fig. S3) revealed that 44% of the total effect is mediated via pathology.

**Table 2.** Effect of *APOE* ε4 genotype on mtDNAcn.

| Region/Study | N of *APOE* ε4 dosages<br>(0, 1, 2) | Unadjusted for pathology | | | Adjusted for pathology* | | |
| --- | --- | --- | --- | --- | --- | --- | --- |
| | | $\beta$ | SE | p | $\beta$ | SE | p |
| DLPFC/ROSMAP | 335, 112, 7 | -0.364 | 0.099 | $2.8 \times 10^{-04}$ | -0.204 | 0.105 | 0.051 |
| CB/ROSMAP | 182, 56, 4 | -0.292 | 0.134 | 0.031 | -0.223 | 0.144 | 0.124 |
| TCX/Mayo | 183, 70, 9 | -0.280 | 0.114 | 0.015 | -0.126 | 0.121 | 0.301 |
| FP/MSBB | 171, 88, 11 | -0.158 | 0.106 | 0.137 | -0.048 | 0.104 | 0.642 |
| Meta-analysis | 871, 326, 31 | -0.275 | 0.056 | $8.0 \times 10^{-07}$ | -0.141 | 0.058 | 0.014 |

\*ROSMAP was adjusted by adding a quantitative score for amyloid and tau burden to the model. Mayo and MSBB were adjusted by adding the pathologic diagnosis to the model.

### mtDNA heteroplasmy levels in the cortex are higher with age

We detected the number of mtDNA heteroplasmic mutations (heteroplasmy level) in the WGS data to assess the relation of mtDNA mutational burden with mtDNAcn and pathologies. Here, we defined a mtDNA heteroplasmic mutation as a point mutation with a relative frequency between 3% and 90%. As in the previous section, we studied the four brain regions with ≥100 samples and excluded the non-Caucasian samples from the MSBB dataset (a total of n=1,228 brain samples). On average, between 2.6 and 2.8 heteroplasmic mutations were observed in the three cortical regions, whereas the CB demonstrated less heteroplasmic mutations (mean of 1.0) (Fig. 4A). As expected, most of the heteroplasmic mutations were located in the mtDNA hypervariable control region, also known as the D-loop (Fig. 4B). None of the genes encoded in the mitochondrial genome showed an enrichment of heteroplasmic mutations.

**Figure 4.**
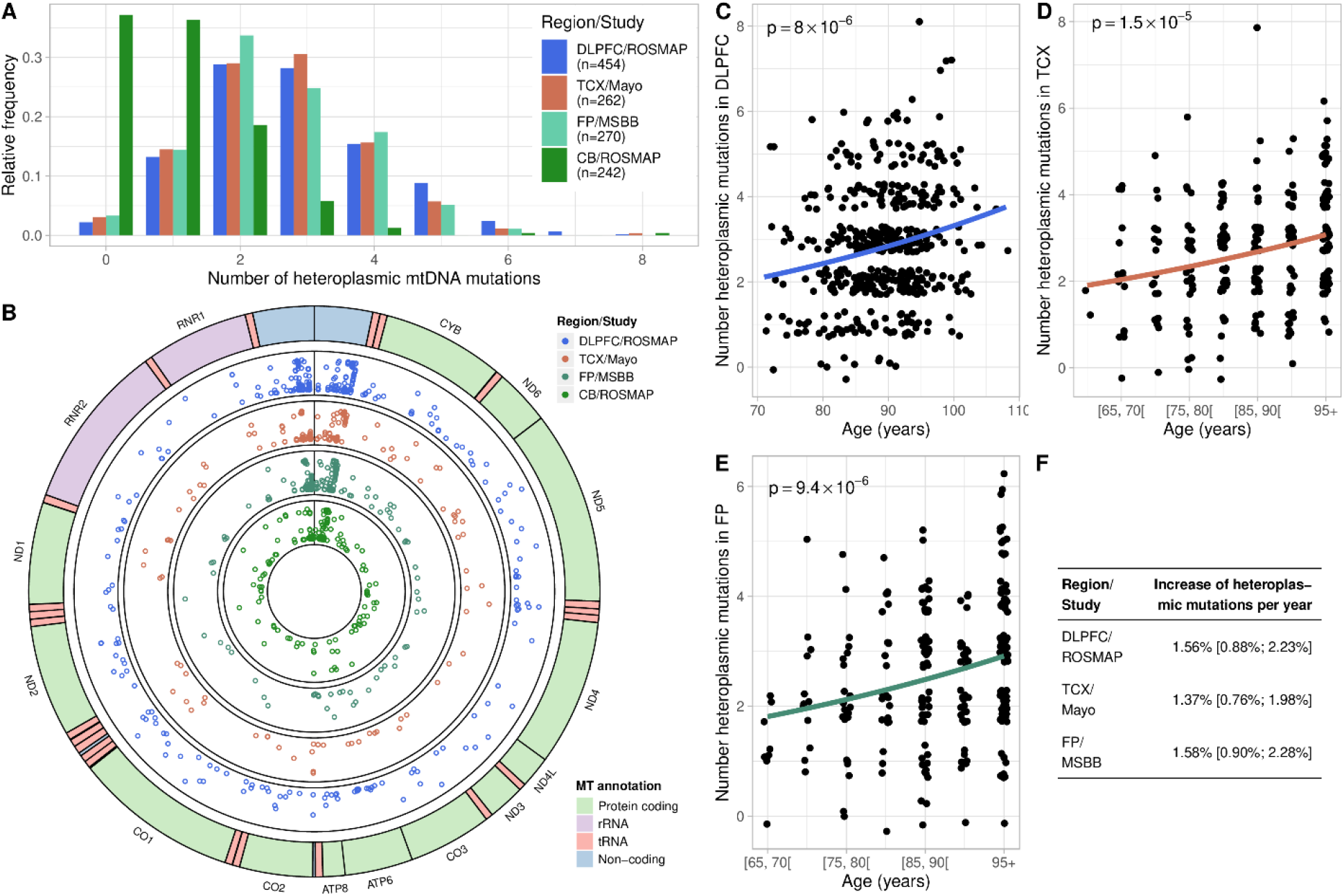
Frequency of mtDNA heteroplasmic mutations in cortical regions increases with age. (A) Histogram depicts the number of mtDNA heteroplasmic mutations detected per sample in each of the four studied brain regions. (B) Circular plot of the mitochondrial genome shows the genome annotation on the outer circle. The four inner circles show the genomic locations and the relative frequencies (y-axis) of mtDNA heterplasmic mutations detected in each of the four brain regions. (C-E) Scatter plots showing the relation between age and mtDNA heteroplasmy burden in the DLPFC (C), in the TCX (D), and in the FP (E). Some jitter was added before plotting the points to avoid overlapping points. Regression curves and p values were obtained from quasi-Poisson regression models with the number of mtDNA heteroplasmic mutations as outcome and age as independent variable. In the TCX and FP, age was grouped into 5-year strata because of censored data. (F) Table shows the effect sizes of age on mtDNA heteroplasmy burden obtained from the Quasi-Poisson regression models depicted in panels (C-E). The 95% confidence intervals are shown in brackets.

Next, we used the DLPFC dataset to study whether the mtDNA heteroplasmy levels are related to brain pathologies, cognitive function, sex, age, and mtDNAcn. In contrast to the mtDNAcn, heteroplasmy levels were not associated with AD pathologies or cognitive function. We also found no significant association between heteroplasmy levels and mtDNAcn in the DLPFC. However, heteroplasmy levels increased significantly with advancing age: we report estimated 1.6% increase per year in the DLPFC (p=8.0 × 10^−6^, quasi-Poisson regression) (Fig. 4C). The accumulation of mtDNA heteroplasmic mutations with aging in the cortex were replicated in the TCX and FP data with similar effect sizes (Fig. 4D-F). In line with the low abundance of heteroplasmic mutations in the CB, the association with age was not detectable in this brain region. The association with age persisted when we adjusted for pathologic diagnosis, sex, and mtDNAcn using multivariable models for each of the three regions DLPFC, TCX, and FP (Fig. S4, Table S5). In addition, we found a weak association with AD diagnosis and with mtDNAcn in the TCX, which were absent in the DLPFC and in the FP. Overall, our analyses showed that age is the primary driver of mtDNA heteroplasmic mutations in the cortical regions and that the CB demonstrates low mtDNA mutation rates.

### Altered mtDNAcn does not necessarily imply alteration in mitochondrial content

A lower mtDNAcn in a cell can reflect a lower mtDNAcn per mitochondrion or a lower mitochondrial content in the cell. We therefore quantified the mitochondrial content in the DLPFC using mass spectrometry-based proteomics data for a subset of 156 subjects who have DLPFC mtDNAcn measurements. Abundances of 10 proteins specific for mitochondria selected from the Human Protein Atlas were quantified and the median of the standardized protein values was used as a mitochondrial content score (Fig. S5A) (Thul et al., 2017). Interestingly, we found no significant correlation between mtDNAcn and mitochondrial content, but both measures were positively correlated with neuronal proportion and cognitive function (Fig. 5A, Fig. S5B). This suggests that these two measures (mtDNAcn and protein-based content) represent different aspects of mitochondrial health that both affect cognition but can change relatively independent from each other. A similar lack of correlation between mtDNAcn and mitochondrial content has been previously reported for muscle tissue (Brinckmann et al., 2010; Larsen et al., 2012). Next, we quantified the abundances of mitochondrial respiratory chain complexes in a similar way using mass spectrometry data from proteins annotated with mitochondrial complex I to V (see methods). Protein levels for the mitochondrial complexes I to V were highly correlated with each other and with mitochondrial content (Fig. 5A). To further disentangle the relationship between mtDNAcn, mitochondrial content, mitochondrial heteroplasmy levels and AD-related phenotypes, we estimated the partial correlations between these variables. The graph in Fig. 5B represents a sparse representation of the partial correlation matrix of the variables. An edge in this graph represents a direct association between two variables that remained when controlling for all other variables in the graph. The stability of the graph’s edges was assessed using bootstrapping (Fig. S5C). The top left of the graph shows age as the primary driver for amyloid-β and mtDNA heteroplasmy levels. The latter are not associated with any other variable in our graph suggesting that mtDNA heteroplasmic mutations are not involved in AD pathogenesis. In the lower part of the graph, amyloid-β is strongly associated with tau, and tau with cognitive function. Tau is also associated with mtDNAcn as suggested by our previous analyses. mtDNAcn and mitochondrial content are not directly connected, but both are associated with neuronal proportion and cognitive function. Together, these findings indicate a complex involvement of mitochondria in neurodegeneration and the need for future studies to measure both parameters to better understand mitochondrial recalibrations in AD.

**Figure 5.**
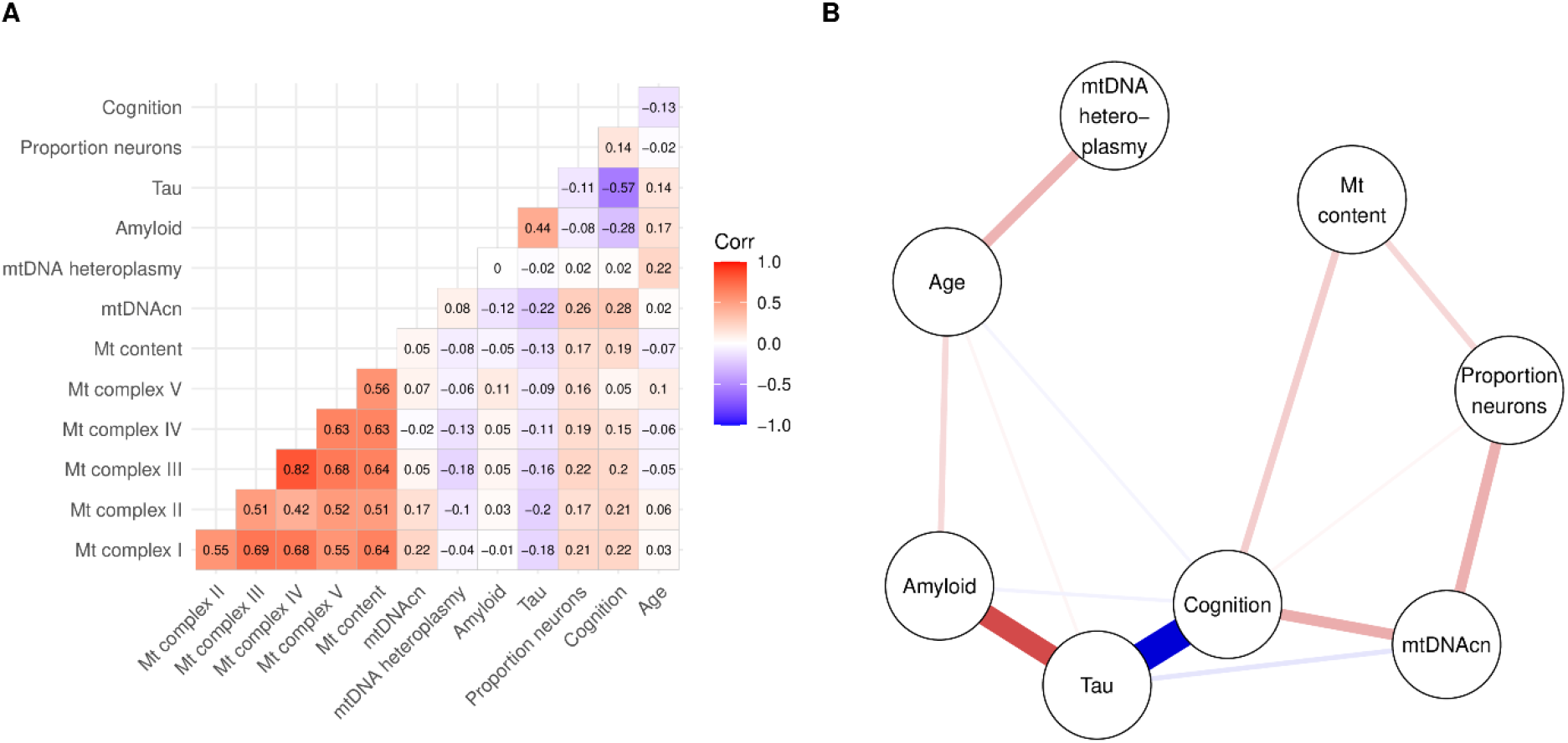
Correlates of mitochondrial health demonstrate complex relationship with AD-related traits in the DLPFC. (A) Heatmap shows pairwise Pearson correlations between mitochondrial complexes I to V scores and mitochondrial content score derived from mass spectrometry data, mtDNAcn and mtDNA heteroplasmy levels derived from WGS data, amyloid and tau derived from immunohistochemistry data, proportion of neurons derived from RNA-seq data, and cognitive functioning and age. Nonparanormal transformation was applied to mtDNA heteroplasmic mutation counts before calculating correlations. Number of available cases for each pair of variables is given in Figure S4B. (B) Graph shows a sparse representation of the partial correlations between the different variables. An edge in this graph indicates that the two connected variables are correlated with each other after controlling for all other variables in the graph. Thickness and color of the edges represent the strength and direction of the partial correlation.

## Discussion

We characterized the mtDNAcn in 1,361 aged human brain samples from five regions and estimated a reduction of mtDNAcn by 7% to 14% in pathologic AD compared to non-AD samples in the cortical regions profiled in this study. We then leveraged the detailed pathologic and cognitive characterization of the ROSMAP study to identify the primary drivers of mtDNAcn loss and to assess the relation to cognitive functioning in the presence of mixed pathologies, which are frequently observed in the aged human brain (Schneider et al., 2007).

In the DLPFC, lower mtDNAcn was primarily associated with tau pathology. When accounting for ten common brain pathologies, tau was the only pathology that remained significantly associated with mtDNAcn. The mtDNAcn at the tissue level depends on the cell type composition of the studied tissue, as is well known in blood (Shim et al., 2020). We therefore estimated the neuronal proportion in the DLPFC samples, showed that the neuronal proportion is associated with mtDNAcn, and confirmed that the association between tau pathology and mtDNAcn was still significant - albeit attenuated - when adjusting for neuronal proportion. Thus, our data suggests that the relationship is not merely caused by tau-driven loss of mitochondria-rich neuronal cells; other mechanisms link tau pathology to reduced mtDNAcn. Mechanisms that could potentially underlie our observation have been explored in model systems. Hyperphosphorylated tau has been shown to impair mitochondrial axonal transport (Kopeikina et al., 2011; Shahpasand et al., 2012) and to affect mitochondrial fission/fusion dynamics (Kandimalla et al., 2016; Li et al., 2016). Tau is also known to be a strong predictor for cognitive decline. Interestingly, when studying cognitive functioning, mtDNAcn was a significant predictor in our model that included tau, nine other brain pathologies, neuronal proportion, and demographic variables. Therefore, improving mitochondrial health in the presence of pathologies might improve cognitive functioning.

In contrast to tau pathology, amyloid-β pathology was not significantly associated with mtDNAcn after accounting for other pathologies. This finding was supported by the results from the Mayo study where the mtDNAcn was not reduced in persons diagnosed with pathologic aging, which is defined by high amyloid-β burden but no or minimal tau pathology (Murray and Dickson, 2014). Interestingly, numerous studies have demonstrated that the amyloid-β precursor protein (APP) as well as amyloid-β peptides localize at mitochondria and affect mitochondrial function and bioenergetics (Devi et al., 2006; Du et al., 2008). Further, C-terminal fragments resulting from the processing of APP (APP-CTFs) have been implicated in AD and may trigger morphological and functional changes of mitochondria (Vaillant-Beuchot et al., 2021). The absence of an association in our data could result from measuring only amyloid-β peptides (1-40) and (1-42) (both were detected by our antibody), which does not fully capture APP processing. Further, amyloid-β could still impair mitochondrial respiratory chain capacity or other non-energetic functions (e.g. calcium handling) without changes in mtDNAcn.

In the PCC, TDP-43 pathology was the most important factor and explained 23% of the mtDNAcn’s variance. The effect of TDP-43 on mitochondria has been mainly studied in model systems for ALS, where suppressing the localization of TDP-43 inside mitochondria reduces TDP-43 toxicity (Wang et al., 2016). When accumulating inside mitochondria, TDP-43 induces the release of mtDNA into the cytoplasm via the permeability transition pore (Yu et al., 2020). Interestingly, the effect of TDP-43 pathology on mtDNAcn was moderate in the DLPFC, which could be caused by the distinct spatial pattern of TDP-43 progression in aged healthy and AD brains, or by a larger susceptibility of the neurons in the PCC to TDP-43 pathology.

The mtDNAcn has been suggested as a biomarker for aging because several studies of peripheral blood have reported an inverse correlation between age and mtDNAcn (Fries et al., 2017; Mengel-From et al., 2014). However, more recent studies demonstrated that the inverse correlation is likely caused by unaccounted age-related changes of cell type proportions in the blood (Moore et al., 2018; Rausser et al., 2021). In our study, we found no association between mtDNAcn and age or sex in the studied brain regions when we adjusted for pathologies. We also found no significant associations in the CB, which accumulates much less AD pathology than the DLPFC or the PCC. In summary, our results indicate that lower mtDNAcn is driven by certain pathologies rather than aging and restricted to brain regions directly affected by the respective pathologies. Similarly, a study of Parkinson’s disease brains found lower mtDNAcn in the vulnerable substantia nigra but not in the frontal cortex, which is less affected in Parkinson’s disease (Pyle et al., 2016).

To assess the effect of genetic variants on the mtDNAcn in the brain, we performed a targeted analysis of 81 lead SNPs that were recently reported by a large GWAS of mtDNAcn in blood (Longchamps et al., 2021). A missense variant in the protease *LONP1* demonstrated a moderate but significant effect size of 0.23 standard deviations larger mtDNAcn per dosage of the alternative allele in our meta-analysis of four brain regions. The original blood-based GWAS also detected a single variant at the *APOE* locus, which harbors the strongest genetic risk factor for AD. We therefore investigated the effect of the *APOE* ε4 haplotype (defined by two coding variants) on the mtDNAcn. While a large fraction of the *APOE* ε4 effect was mediated via AD pathologies, we also found evidence for a direct effect on the mtDNAcn. These findings support the hypothesis that the *APOE* ε4 allele exerts its risk not only via regulating amyloid-β aggregation and clearance but also through other pathways, including mitochondrial bioenergetics (Huang, 2010; Liu et al., 2013). In summary, while this study is the first, to our knowledge, to report evidence for genetic regulation of the mtDNAcn in the brain, the sample size was a limiting factor and future non-targeted studies with much larger sample sizes will likely detect more loci.

Several tissues in addition to brain accumulate mtDNA mutations during aging (Li et al., 2015; Lin et al., 2002; Zhang et al., 2017). Here, we found higher heteroplasmy levels with age in three cortical regions and estimated an association consistent with accumulation rate of about 1.5% per year in this age group. We did not find a significant association with age in the CB where the overall frequency of mtDNA heteroplasmic mutations was very low (mean of 1.0) supporting the theory of region-specific accumulation of mtDNA mutations in the brain (Keogh and Chinnery, 2015). Consistently, a previous study (Wei et al., 2017) failed to detect an association between age and heteroplasmy levels in their samples which were mainly (87%) obtained from the CB. Thus, the CB seems to acquire less heteroplasmic mutations than the cortex or the heteroplasmic mutations in the CB have, on average, a lower relative frequency which may often not surpass the detection threshold of 3% used in this study. In contrast to the mtDNAcn, we found little evidence for the involvement of mtDNA heteroplasmy in AD or in the development of other brain pathologies. Further, mtDNA heteroplasmy levels were not related to the mtDNAcn. Overall, our work showed that mtDNA heteroplasmic mutations accumulate over an individual’s life-time in the cortex but are not related to neurodegenerative diseases. The seeming insignificance of low-level mtDNA heteroplasmy is consistent with a large reserve respiratory capacity, such that mtDNAcn is generally in excess (>50%) of the minimum number of mtDNA copies required to sustain bioenergetic capacity (Rossignol et al., 2003).

A lower mtDNAcn can be caused by lower mitochondrial mass in the cells or by a lower mtDNAcn per mitochondrion. We generated a proteomic measure of mitochondrial content for a subset of our DLPFC samples and showed that the mtDNAcn and mitochondrial content vary relatively independent of each other in the DLPFC. The decoupling of the two variables may be explained by distinct mechanisms that regulate mtDNAcn and mitochondrial content and that have been successfully manipulated in model systems to regulate one of the readouts, without affecting the other one (Filograna et al., 2020). Further, mitochondrial content but not mtDNAcn was correlated with the protein scores for the respiratory chain complexes, suggesting that the mtDNAcn does probably not directly reflect respiratory capacity. When we integrated the detailed pathologic and cognitive measures with the mitochondrial measures available for our DLPFC samples to disentangle their relationship, we found that both, mtDNAcn and mitochondrial content, were associated with cognitive functioning and neuronal loss indicating their involvement in neurodegeneration and the need for future studies to interrogate several mitochondrial measures to fully capture mitochondrial health.

## Methods

### ROSMAP cohort and neuropathologic characterization

The Religious Orders Study (ROS) and the Rush Memory and Aging Project (MAP) are two cohort studies of aging and dementia conducted by the same team of investigators and share a large common core of harmonized clinical and post mortem data collection which allows for joint analyses (Bennett et al., 2018). Participants entered the studies without known dementia and agreed to annual clinical and cognitive assessments as well as brain donation after death. Both cohort studies (ROS and MAP) were approved by an Institutional Review Board of Rush University Medical Center. All participants signed an informed consent, an Anatomic Gift Act for brain donation, and a repository consent to allow their data and biospecimens to be shared. More information on the study and resources can be found on our Resource Sharing Hub at https://www.radc.rush.edu.

To obtain a pathologic diagnosis of AD, a modified Bielschowsky silver stain was used to visualize neuritic plaques, diffuse plaques, and neurofibrillary tangles in five cortical regions (hippocampus, entorhinal, midfrontal, middle temporal, and inferior parietal). A board-certified neuropathologist, blinded to clinical data, determined the pathologic diagnosis of AD based on an intermediate to high likelihood of AD according to the NIA Reagan criteria. The quantitative global AD pathology score was derived from the Bielschowsky silver stains by counting and standardizing each of the pathologies neuritic plaques, diffuse plaques, and neurofibrillary tangles in each of the five cortical regions, and then averaging the standardized measures across regions. The average of these three pathology measures was then used as single score of AD pathology burden. Amyloid-β and tau tangles were assessed in 8 brain regions (hippocampus, entorhinal cortex, midfrontal cortex, inferior temporal gyrus, angular gyrus, calcarine cortex, anterior cingulate cortex, and superior frontal cortex) using immunochemistry (Bennett et al., 2006; Bennett et al., 2004). Paraffin-embedded sections were immunostained for amyloid-β using 1 of 3 monoclonal anti-human antibodies: 4G8 (1:9000; Covance Labs, Madison, WI), 6F/3D (1:50; Dako North America Inc., Carpinteria, CA), and 10D5 (1:600; Elan Pharmaceuticals, San Francisco, CA). Paired helical filament (PHF) tau tangles were labeled with an antibody specific to phosphorylated tau (AT8, Thermoscientific, Waltham, MA, USA). A computerized sampling procedure combined with image analysis software was used to calculate the percentage area occupied with amyloid-β and the density of PHFtau tangles. Composite scores were computed for overall amyloid-β burden and tau tangle density by averaging the scores obtained from the eight brain regions. The three quantitative measurements (global AD pathology, amyloid-β, and tau) were square root transformed for better statistical properties.

Presence of TDP-43 cytoplasmic inclusions in neurons and glia were determined in eight regions (amygdala, entorhinal cortex, hippocampus CA1, hippocampus dentate gyrus, anterior temporal pole cortex, midtemporal cortex, orbital frontal cortex, and midfrontal cortex) using a phosphorylated monoclonal TAR5P-1D3 (pS409/410; 1:100, Ascenion, Munich, Germany) TDP-43 antibody. Based on the absence or presence of TDP-43 pathology in the eight regions, four stages of TDP-43 distribution were recognized: none, amygdala, amygdala + limbic, amygdala + limbic + neocortical (Nag et al., 2017). Lewy body disease was assessed in four stages (not present, nigral-predominant, limbic-type, neocortical-type). Seven regions (substantia nigra, anterior cingulate cortex, entorhinal cortex, amygdala, midfrontal cortex, superior or middle temporal cortex, inferior parietal cortex) were assessed for Lewy bodies using α-synuclein immunostaining (LB509; 1:150 or 1:100, Zymed Labs, Invitrogen, Carlsbad, CA, USA; and pSyn#64; 1:20,000; Wako Chemicals, Richmond, VA,USA) (Schneider et al., 2012). Cerebral amyloid angiopathy pathology was assessed in four neocortical regions (midfrontal, midtemporal, parietal, and calcarine cortices) using using 1 of 3 monoclonal anti-human antibodies: 4G8 (1:9000; Covance Labs, Madison, WI), 6F/3D (1:50; Dako North America Inc., Carpinteria, CA), and 10D5 (1:600; Elan Pharmaceuticals, San Francisco, CA). Similar to the protocol by Love et al. (2014), meningeal and parenchymal vessels were assessed for amyloid-β deposition in each region and scored from 0 to 4. Scores were averaged across the four regions and categorized into none, mild, moderate, or severe (Boyle et al., 2015). Large vessel cerebral atherosclerosis rating was made by visual inspection after paraformaldehyde fixation, at the Circle of Willis at the base of the brain, and included evaluation of the vertebral, basilar, posterior cerebral, middle cerebral, and anterior cerebral arteries and their proximal branches. Severity was graded (none or possible, mild, moderate, severe) by visual examination of the extent of involvement of each artery and number of arteries involved (Arvanitakis et al., 2017). Arteriolosclerosis was graded by evaluating the vessels of the anterior basal ganglia for histological changes as previously described (Buchman et al., 2011). Four stages (none, mild, moderate, and severe) were recognized. The presence of one or more gross chronic cerebral infarctions was determined during gross examination and confirmed histologically. The presence of one or more chronic microinfarcts was determined on sections of a minimum of nine regions stained with hematoxylin and eosin (H&E) (Arvanitakis et al., 2011). The presence of hippocampal sclerosis was identified by severe neuronal loss and gliosis on H&E-stained sections in CA1 or subiculum (Nag et al., 2015).

DNA for WGS was extracted from the DLPFC using Qiagen’s QIAamp DNA kit (n=367) or Qiagen’s AllPrep Universal kit (n=87), from the PCC using Qiagen’s AllPrep Universal kit (n=66), or from the CB using Qiagen’s Gentra Puregene Tissue kit (n=242). Sample characteristics are summarized in Table 1.

### Mayo and MSSB cohorts

Samples included in the Mayo case-control study were obtained either from the Mayo Brain Bank or from the Banner Sun Health Institute and classified as control, AD, PSP or pathologic aging based on neuropathological assessment (Allen et al., 2016). The study was approved by the Mayo Clinic Institutional Review Board. All AD samples demonstrated tau pathology (Braak score ≥ 4). All controls demonstrated no or minimal tau pathology (Braak score ≤ 3) and were without any other neurodegenerative disease. Here, samples from the Banner Sun Health Institute were excluded since all AD samples were obtained from the Mayo Brain Bank and we observed a difference in the mtDNAcn between control samples from the two different centers. Since age at death was right-censored at 95 years for HIPPA compliance, we stratified age into 5 year strata with an open interval > 95 years for our analyses. Specimens for WGS were sampled from the temporal cortex (n=262). Sample characteristics are summarized in Table S1.

The MSBB case-control study cohort was assembled after applying stringent inclusion/exclusion criteria and represents the full spectrum of cognitive and neuropathological AD severity in the absence of discernable non-AD neuropathology (Wang et al., 2018). The study was approved by the Mount Sinai and JJ Peters VA Medical Center Institutional Review Boards. Neuropathological assessments were performed according to the Consortium to Establish a Registry for Alzheimer’s Disease (CERAD) protocol and included assessment by hematoxylin and eosin, modified Bielschowski, modified thioflavin S, and anti-amyloid-β (4G8), anti-tau (AD2) and anti-ubiquitin (Dakoa Corp.). Pathologic AD was defined based on the CERAD stages definitive AD and probably AD. Samples staged as possible AD or no AD were considered as controls. Age at death was right-censored and stratified as for the Mayo cohort.

Specimens for WGS were sampled from the frontal pole (n=337). Sample characteristics are summarized in Table S2. A total of n=67 samples were obtained from non-Caucasians and excluded from the mtDNAcn GWAS and mtDNA heteroplasmy analyses.

### Whole-genome sequencing

WGS libraries from all three studies were prepared using the KAPA Hyper Library Preparation Kit in accordance with the manufacturer’s instructions. Briefly, 650ng of DNA was sheared using a Covaris LE220 sonicator (adaptive focused acoustics). DNA fragments underwent bead-based size selection and were subsequently end-repaired, adenylated, and ligated to Illumina sequencing adapters. Final libraries were evaluated using fluorescent-based assays including qPCR with the Universal KAPA Library Quantification Kit and Fragment Analyzer (Advanced Analytics) or BioAnalyzer (Agilent 2100). Libraries were sequenced on an Illumina HiSeq X sequencer (v2.5 chemistry) using 2 × 150bp cycles.

### Variant calling

Sequence alignment and nuclear DNA variant calling was performed by the automated pipeline of the New York Genome Center, where all samples were sequenced (De Jager et al., 2018). Briefly, paired-end 150 bp reads were aligned to the GRCh37 human reference using the Burrows-Wheeler Aligner (BWA-MEM v0.7.8) and processed using the GATK best-practices workflow that includes marking of duplicate reads using Picard tools v1.83, local realignment around indels, and base quality score recalibration (BQSR) using the Genome Analysis Toolkit (GATK v3.4.0).

Mitochondrial homo- and heteroplasmic variants were called following GATK’s Best Practices Mitochondria Pipeline 1.1.0 (https://github.com/gatk-workflows/gatk4-mitochondria-pipeline). GATK v4.1.2 was used to run the pipeline with the rCRS (NC_012920.1) as mtDNA reference sequence. Briefly, sequence reads (mapped to MT or unmapped in original bam file) were mapped to the rCRS and to the rCRS shifted by 8,000 base pairs using BWA-MEM v0.7.8. Variants were detected using GATK’s Mutect2 in both bam files. Subsequently, variants were merged into one VCF file using the shifted rCRS for variants around the artificial start/end position of the circular genome and the unmodified rCRS for the remaining part the MT genome. Variant filtering included the median autosomal chromosome coverage to filter potential polymorphic NuMT variants, the mtDNA contamination estimated by the haplochecker (mitolib 0.1.2) to account for possible contamination, a minimum minor allele frequency of 0.03, and an F-score beta of 1 (default settings). Bam and VCF are available at the AD Knowledge Portal (see data availability section).

### Estimation of the mtDNAcn from WGS data

The median sequence coverages of the autosomal chromosomes *cov*_*nuc*_ and of the mitochondrial genome *cov*_*mt*_ were calculated using R/Bioconductor (packages GenomicAlignments and GenomicRanges). Ambiguous regions were excluded using the intra-contig ambiguity mask from the BSgenome package. The mtDNAcn was defined as (*cov*_*mt*_/*cov*_*nuc*_) × 2. Raw mtDNAcn was used for the first analysis shown in Fig. 1. For subsequent analyses the mtDNAcn was z-standardized within each brain region and DNA extraction kit and then logarithmized. The normalization facilitated the combined analysis of the two different kits used for the DLPFC and resulted in approximately normal mtDNAcn measures (Fig. S1).

### Estimation of the neuronal proportion from RNA-seq data

Proportion of neurons were estimated for n=327 DLPFC samples with RNA-seq data by applying the Digital Sorting Algorithm (DSA) (Zhong et al., 2013) to published marker genes that were previously used to deconvolute cortical RNA-seq data (Wang et al., 2020). RNA-seq data were TMM normalized and technical variables were regressed out. Only marker genes with a mean transcription level ≥2 cpm in our dataset were used (87 marker for astrocytes, 88 for endothelial cells, 59 for microglia, 90 for neurons, and 86 for oligodendrocytes). As proposed by Wang et al. (2020), DSA was modified so that the median instead of the mean transcription level of all marker genes per cell type was calculated.

### Estimation of mitochondrial content from proteomic data

Tandem mass tag (TMT) multiplexed mass spectrometry data was available for a subset of n=156 DLPFC samples that also had WGS data. Mass spectrometry data was preprocessed and normalized as previously described (Wingo et al., 2020). Ten proteins were selected to quantify mitochondrial mass. Six proteins (CS, LRPPRC, SLC25A24, TIMM44, GCDH, and TRAP1) were taken from the Human Protein Atlas’ list of mitochondrial marker proteins (Thul et al., 2017). The four additional proteins (HSPD1, VDAC2, VDAC3, and TOMM20) have been previously used as mitochondrial markers and confirmed to be specific to mitochondria in the Human Protein Atlas. The median protein level of these ten proteins was used as mitochondrial content measure. Fig. S5A shows the correlation between the 10 selected proteins. Mitochondrial complexes I-V were quantified by calculating the median of all proteins that were annotated by the respective GO cellular component term.

### Statistical methods

Statistical analyses were conducted in R. Standard multivariable linear regression models were used when the standardized mtDNAcn or cognitive function were the dependent variable. Quasi-Poisson regression models were used when the number of mtDNA heteroplasmic mutations (heteroplasmy level) was the dependent variable. Reported p values were obtained from likelihood ratio tests and unadjusted unless stated otherwise in the text. Continuous variables were standardized to obtain comparable effect sizes unless stated otherwise. All variables included in the respective models are described in the text and figures. When analyzing the effect of SNPs on mtDNAcn, population structure was modelled by the first three principle components obtained from the genotypes after pruning. Random effects meta-analyses were performed using the R package meta v4.13. P values and quasi-Bayesian confidence intervals for the mediation analysis were calculated using the R package mediation v4.5.0 (Imai et al., 2010). The sparse Gaussian graphical model (Fig. 5B) was estimated by the graphical LASSO method with EBIC model selection and a default tuning parameter of 0.5 as implemented in the R package bootnet (Epskamp et al., 2018). The nonparanormal transformation implemented in the package huge was applied to relax the assumption of normality for the mtDNA heteroplasmy levels before estimating the network. Network stability was assessed using 1,000 bootstraps.

## Data Availability

Raw and processed data (WGS, proteomics, RNA-seq), called variants, and analysis output (mtDNA, mtDNA heteroplasmy levels) from the three studies (see Table S6 for details) are available via the AD Knowledge Portal (https://adknowledgeportal.org). The AD Knowledge Portal is a platform for accessing data, analyses, and tools generated by the Accelerating Medicines Partnership (AMP-AD) Target Discovery Program and other National Institute on Aging (NIA)-supported programs to enable open-science practices and accelerate translational learning. The data, analyses and tools are shared early in the research cycle without a publication embargo on secondary use. Data is available for general research use according to the following requirements for data access and data attribution (https://adknowledgeportal.org/DataAccess/Instructions).
For access to content described in this manuscript see: https://doi.org/10.7303/syn25618990
ROSMAP pathologic and phenotypic data are available at https://www.radc.rush.edu.
R and shell scripts used to generate figures and analysis results are deposited at GitHub: https://github.com/cu-ctcn/mtDNA

## Acknowledgement

This work was supported by NIH grants U01AG046152 (PLD, DAB), U01AG061356 (PLD, DAB), P30AG010161 (DAB), R01AG015819 (DAB), and R01AG017917 (DAB).

## Data availability

Raw and processed data (WGS, proteomics, RNA-seq), called variants, and analysis output (mtDNA, mtDNA heteroplasmy levels) from the three studies (see Table S6 for details) are available via the AD Knowledge Portal (https://adknowledgeportal.org). The AD Knowledge Portal is a platform for accessing data, analyses, and tools generated by the Accelerating Medicines Partnership (AMP-AD) Target Discovery Program and other National Institute on Aging (NIA)-supported programs to enable open-science practices and accelerate translational learning. The data, analyses and tools are shared early in the research cycle without a publication embargo on secondary use. Data is available for general research use according to the following requirements for data access and data attribution (https://adknowledgeportal.org/DataAccess/Instructions).

For access to content described in this manuscript see: https://doi.org/10.7303/syn25618990 ROSMAP pathologic and phenotypic data are available at https://www.radc.rush.edu.

R and shell scripts used to generate figures and analysis results are deposited at GitHub: https://github.com/cu-ctcn/mtDNA

## References

Allen, M., Carrasquillo, M.M., Funk, C., Heavner, B.D., Zou, F., Younkin, C.S., Burgess, J.D., Chai, H.S., Crook, J., Eddy, J.A., et al. (2016). Human whole genome genotype and transcriptome data for Alzheimer’s and other neurodegenerative diseases. Sci Data 3, 160089.

Area-Gomez, E., Guardia-Laguarta, C., Schon, E.A., and Przedborski, S. (2019). Mitochondria, OxPhos, and neurodegeneration: cells are not just running out of gas. J Clin Invest 129, 34–45.

Area-Gomez, E., Larrea, D., Pera, M., Agrawal, R.R., Guilfoyle, D.N., Pirhaji, L., Shannon, K., Arain, H.A., Ashok, A., Chen, Q., et al. (2020). APOE4 is Associated with Differential Regional Vulnerability to Bioenergetic Deficits in Aged APOE Mice. Sci Rep 10, 4277.

Arvanitakis, Z., Capuano, A.W., Leurgans, S.E., Buchman, A.S., Bennett, D.A., and Schneider, J.A. (2017). The Relationship of Cerebral Vessel Pathology to Brain Microinfarcts. Brain Pathol 27, 77–85.

Arvanitakis, Z., Leurgans, S.E., Barnes, L.L., Bennett, D.A., and Schneider, J.A. (2011). Microinfarct pathology, dementia, and cognitive systems. Stroke 42, 722–727.

Bender, A., Krishnan, K.J., Morris, C.M., Taylor, G.A., Reeve, A.K., Perry, R.H., Jaros, E., Hersheson, J.S., Betts, J., Klopstock, T., et al. (2006). High levels of mitochondrial DNA deletions in substantia nigra neurons in aging and Parkinson disease. Nat Genet 38, 515–517.

Bennett, D.A., Buchman, A.S., Boyle, P.A., Barnes, L.L., Wilson, R.S., and Schneider, J.A. (2018). Religious Orders Study and Rush Memory and Aging Project. J Alzheimers Dis 64, S161–S189.

Bennett, D.A., Schneider, J.A., Tang, Y., Arnold, S.E., and Wilson, R.S. (2006). The effect of social networks on the relation between Alzheimer’s disease pathology and level of cognitive function in old people: a longitudinal cohort study. Lancet Neurol 5, 406–412.

Bennett, D.A., Schneider, J.A., Wilson, R.S., Bienias, J.L., and Arnold, S.E. (2004). Neurofibrillary tangles mediate the association of amyloid load with clinical Alzheimer disease and level of cognitive function. Arch Neurol 61, 378–384.

Bota, D.A., and Davies, K.J. (2016). Mitochondrial Lon protease in human disease and aging: Including an etiologic classification of Lon-related diseases and disorders. Free Radic Biol Med 100, 188–198.

Botha, H., Mantyh, W.G., Murray, M.E., Knopman, D.S., Przybelski, S.A., Wiste, H.J., Graff-Radford, J., Josephs, K.A., Schwarz, C.G., Kremers, W.K., et al. (2018). FDG-PET in tau-negative amnestic dementia resembles that of autopsy-proven hippocampal sclerosis. Brain 141, 1201–1217.

Boyle, P.A., Yu, L., Nag, S., Leurgans, S., Wilson, R.S., Bennett, D.A., and Schneider, J.A. (2015). Cerebral amyloid angiopathy and cognitive outcomes in community-based older persons. Neurology 85, 1930–1936.

Brinckmann, A., Weiss, C., Wilbert, F., von Moers, A., Zwirner, A., Stoltenburg-Didinger, G., Wilichowski, E., and Schuelke, M. (2010). Regionalized pathology correlates with augmentation of mtDNA copy numbers in a patient with myoclonic epilepsy with ragged-red fibers (MERRF-syndrome). PLoS One 5, e13513.

Buchman, A.S., Leurgans, S.E., Nag, S., Bennett, D.A., and Schneider, J.A. (2011). Cerebrovascular disease pathology and parkinsonian signs in old age. Stroke 42, 3183–3189.

Coskun, P.E., Beal, M.F., and Wallace, D.C. (2004). Alzheimer’s brains harbor somatic mtDNA control-region mutations that suppress mitochondrial transcription and replication. Proc Natl Acad Sci U S A 101, 10726–10731.

D’Erchia, A.M., Atlante, A., Gadaleta, G., Pavesi, G., Chiara, M., De Virgilio, C., Manzari, C., Mastropasqua, F., Prazzoli, G.M., Picardi, E., et al. (2015). Tissue-specific mtDNA abundance from exome data and its correlation with mitochondrial transcription, mass and respiratory activity. Mitochondrion 20, 13–21.

De Jager, P.L., Ma, Y., McCabe, C., Xu, J., Vardarajan, B.N., Felsky, D., Klein, H.U., White, C.C., Peters, M.A., Lodgson, B., et al. (2018). A multi-omic atlas of the human frontal cortex for aging and Alzheimer’s disease research. Sci Data 5, 180142.

Devi, L., Prabhu, B.M., Galati, D.F., Avadhani, N.G., and Anandatheerthavarada, H.K. (2006). Accumulation of amyloid precursor protein in the mitochondrial import channels of human Alzheimer’s disease brain is associated with mitochondrial dysfunction. J Neurosci 26, 9057–9068.

Ding, J., Sidore, C., Butler, T.J., Wing, M.K., Qian, Y., Meirelles, O., Busonero, F., Tsoi, L.C., Maschio, A., Angius, A., et al. (2015). Assessing Mitochondrial DNA Variation and Copy Number in Lymphocytes of ∼2,000 Sardinians Using Tailored Sequencing Analysis Tools. PLoS Genet 11, e1005306.

Du, H., Guo, L., Fang, F., Chen, D., Sosunov, A.A., McKhann, G.M., Yan, Y., Wang, C., Zhang, H., Molkentin, J.D., et al. (2008). Cyclophilin D deficiency attenuates mitochondrial and neuronal perturbation and ameliorates learning and memory in Alzheimer’s disease. Nat Med 14, 1097–1105.

Epskamp, S., Borsboom, D., and Fried, E.I. (2018). Estimating psychological networks and their accuracy: A tutorial paper. Behavior Research Methods 50, 195–212.

Filograna, R., Mennuni, M., Alsina, D., and Larsson, N.G. (2020). Mitochondrial DNA copy number in human disease: the more the better? FEBS Lett.

Frahm, T., Mohamed, S.A., Bruse, P., Gemund, C., Oehmichen, M., and Meissner, C. (2005). Lack of age-related increase of mitochondrial DNA amount in brain, skeletal muscle and human heart. Mech Ageing Dev 126, 1192–1200.

Fries, G.R., Bauer, I.E., Scaini, G., Wu, M.J., Kazimi, I.F., Valvassori, S.S., Zunta-Soares, G., Walss-Bass, C., Soares, J.C., and Quevedo, J. (2017). Accelerated epigenetic aging and mitochondrial DNA copy number in bipolar disorder. Transl Psychiatry 7, 1283.

Guo, W., Jiang, L., Bhasin, S., Khan, S.M., and Swerdlow, R.H. (2009). DNA extraction procedures meaningfully influence qPCR-based mtDNA copy number determination. Mitochondrion 9, 261–265.

Hagg, S., Jylhava, J., Wang, Y., Czene, K., and Grassmann, F. (2020). Deciphering the genetic and epidemiological landscape of mitochondrial DNA abundance. Hum Genet.

Huang, Y. (2010). Abeta-independent roles of apolipoprotein E4 in the pathogenesis of Alzheimer’s disease. Trends Mol Med 16, 287–294.

Imai, K., Keele, L., and Yamamoto, T. (2010). Identification, Inference and Sensitivity Analysis for Causal Mediation Effects. Statistical Science 25, 51–71.

Josephs, K.A., Murray, M.E., Whitwell, J.L., Tosakulwong, N., Weigand, S.D., Petrucelli, L., Liesinger, A.M., Petersen, R.C., Parisi, J.E., and Dickson, D.W. (2016). Updated TDP-43 in Alzheimer’s disease staging scheme. Acta Neuropathol 131, 571–585.

Kandimalla, R., Manczak, M., Fry, D., Suneetha, Y., Sesaki, H., and Reddy, P.H. (2016). Reduced dynamin-related protein 1 protects against phosphorylated Tau-induced mitochondrial dysfunction and synaptic damage in Alzheimer’s disease. Hum Mol Genet 25, 4881–4897.

Keogh, M.J., and Chinnery, P.F. (2015). Mitochondrial DNA mutations in neurodegeneration. Biochim Biophys Acta 1847, 1401–1411.

Kopeikina, K.J., Carlson, G.A., Pitstick, R., Ludvigson, A.E., Peters, A., Luebke, J.I., Koffie, R.M., Frosch, M.P., Hyman, B.T., and Spires-Jones, T.L. (2011). Tau accumulation causes mitochondrial distribution deficits in neurons in a mouse model of tauopathy and in human Alzheimer’s disease brain. Am J Pathol 179, 2071–2082.

Larsen, S., Nielsen, J., Hansen, C.N., Nielsen, L.B., Wibrand, F., Stride, N., Schroder, H.D., Boushel, R., Helge, J.W., Dela, F., et al. (2012). Biomarkers of mitochondrial content in skeletal muscle of healthy young human subjects. J Physiol 590, 3349–3360.

Larsson, N.G. (2010). Somatic mitochondrial DNA mutations in mammalian aging. Annu Rev Biochem 79, 683–706.

Li, M., Schroder, R., Ni, S., Madea, B., and Stoneking, M. (2015). Extensive tissue-related and allele-related mtDNA heteroplasmy suggests positive selection for somatic mutations. Proc Natl Acad Sci U S A 112, 2491–2496.

Li, X.C., Hu, Y., Wang, Z.H., Luo, Y., Zhang, Y., Liu, X.P., Feng, Q., Wang, Q., Ye, K., Liu, G.P., et al. (2016). Human wild-type full-length tau accumulation disrupts mitochondrial dynamics and the functions via increasing mitofusins. Sci Rep 6, 24756.

Lin, M.T., Simon, D.K., Ahn, C.H., Kim, L.M., and Beal, M.F. (2002). High aggregate burden of somatic mtDNA point mutations in aging and Alzheimer’s disease brain. Hum Mol Genet 11, 133–145.

Liu, C.C., Liu, C.C., Kanekiyo, T., Xu, H., and Bu, G. (2013). Apolipoprotein E and Alzheimer disease: risk, mechanisms and therapy. Nat Rev Neurol 9, 106–118.

Longchamps, R., Yang, S., Castellani, C., Shi, W., Lane, J., Grove, M., Bartz, T., Sarnowski, C., Burrows, K., Guyatt, A., et al. (2021). Genome-wide analysis of mitochondrial DNA copy number reveals multiple loci implicated in nucleotide metabolism, platelet activation, and megakaryocyte proliferation. bioRxiv.

Longchamps, R.J., Castellani, C.A., Yang, S.Y., Newcomb, C.E., Sumpter, J.A., Lane, J., Grove, M.L., Guallar, E., Pankratz, N., Taylor, K.D., et al. (2020). Evaluation of mitochondrial DNA copy number estimation techniques. PLoS One 15, e0228166.

Love, S., Chalmers, K., Ince, P., Esiri, M., Attems, J., Jellinger, K., Yamada, M., McCarron, M., Minett, T., Matthews, F., et al. (2014). Development, appraisal, validation and implementation of a consensus protocol for the assessment of cerebral amyloid angiopathy in post-mortem brain tissue. Am J Neurodegener Dis 3, 19–32.

Mengel-From, J., Thinggaard, M., Dalgard, C., Kyvik, K.O., Christensen, K., and Christiansen, L. (2014). Mitochondrial DNA copy number in peripheral blood cells declines with age and is associated with general health among elderly. Hum Genet 133, 1149–1159.

Miller, F.J., Rosenfeldt, F.L., Zhang, C., Linnane, A.W., and Nagley, P. (2003). Precise determination of mitochondrial DNA copy number in human skeletal and cardiac muscle by a PCR-based assay: lack of change of copy number with age. Nucleic Acids Res 31, e61.

Moore, A.Z., Ding, J., Tuke, M.A., Wood, A.R., Bandinelli, S., Frayling, T.M., and Ferrucci, L. (2018). Influence of cell distribution and diabetes status on the association between mitochondrial DNA copy number and aging phenotypes in the InCHIANTI study. Aging Cell 17.

Murray, M.E., and Dickson, D.W. (2014). Is pathological aging a successful resistance against amyloid-beta or preclinical Alzheimer’s disease? xsAlzheimers Res Ther 6, 24.

Nacheva, E., Mokretar, K., Soenmez, A., Pittman, A.M., Grace, C., Valli, R., Ejaz, A., Vattathil, S., Maserati, E., Houlden, H., et al. (2017). DNA isolation protocol effects on nuclear DNA analysis by microarrays, droplet digital PCR, and whole genome sequencing, and on mitochondrial DNA copy number estimation. PLoS One 12, e0180467.

Nag, S., Yu, L., Boyle, P.A., Leurgans, S.E., Bennett, D.A., and Schneider, J.A. (2018). TDP-43 pathology in anterior temporal pole cortex in aging and Alzheimer’s disease. Acta Neuropathol Commun 6, 33.

Nag, S., Yu, L., Capuano, A.W., Wilson, R.S., Leurgans, S.E., Bennett, D.A., and Schneider, J.A. (2015). Hippocampal sclerosis and TDP-43 pathology in aging and Alzheimer disease. Ann Neurol 77, 942–952.

Nag, S., Yu, L., Wilson, R.S., Chen, E.Y., Bennett, D.A., and Schneider, J.A. (2017). TDP-43 pathology and memory impairment in elders without pathologic diagnoses of AD or FTLD. Neurology 88, 653–660.

Nelson, P.T., Dickson, D.W., Trojanowski, J.Q., Jack, C.R., Boyle, P.A., Arfanakis, K., Rademakers, R., Alafuzoff, I., Attems, J., Brayne, C., et al. (2019). Limbic-predominant age-related TDP-43 encephalopathy (LATE): consensus working group report. Brain 142, 1503–1527.

Nunnari, J., and Suomalainen, A. (2012). Mitochondria: in sickness and in health. Cell 148, 1145–1159.

Pyle, A., Anugrha, H., Kurzawa-Akanbi, M., Yarnall, A., Burn, D., and Hudson, G. (2016). Reduced mitochondrial DNA copy number is a biomarker of Parkinson’s disease. Neurobiol Aging 38, 216 e217–216 e210.

Rausser, S., Trumpff, C., McGill, M.A., Junker, A., Wang, W., Ho, S., Mitchell, A., Karan, K.R., Monk, C., Segerstrom, S.C., et al. (2021). Mitochondrial phenotypes in purified human immune cell subtypes and cell mixtures. bioRxiv, 2020.2010.2016.342923.

Rice, A.C., Keeney, P.M., Algarzae, N.K., Ladd, A.C., Thomas, R.R., and Bennett, J.P., Jr. (2014). Mitochondrial DNA copy numbers in pyramidal neurons are decreased and mitochondrial biogenesis transcriptome signaling is disrupted in Alzheimer’s disease hippocampi. J Alzheimers Dis 40, 319–330.

Rodriguez-Santiago, B., Casademont, J., and Nunes, V. (2001). Is mitochondrial DNA depletion involved in Alzheimer’s disease? Eur J Hum Genet 9, 279–285.

Rossignol, R., Faustin, B., Rocher, C., Malgat, M., Mazat, J.P., and Letellier, T. (2003). Mitochondrial threshold effects. Biochem J 370, 751–762.

Schneider, J.A., Arvanitakis, Z., Bang, W., and Bennett, D.A. (2007). Mixed brain pathologies account for most dementia cases in community-dwelling older persons. Neurology 69, 2197–2204.

Schneider, J.A., Arvanitakis, Z., Yu, L., Boyle, P.A., Leurgans, S.E., and Bennett, D.A. (2012). Cognitive impairment, decline and fluctuations in older community-dwelling subjects with Lewy bodies. Brain 135, 3005–3014.

Shahpasand, K., Uemura, I., Saito, T., Asano, T., Hata, K., Shibata, K., Toyoshima, Y., Hasegawa, M., and Hisanaga, S. (2012). Regulation of mitochondrial transport and inter-microtubule spacing by tau phosphorylation at the sites hyperphosphorylated in Alzheimer’s disease. J Neurosci 32, 2430–2441.

Shim, H.B., Arshad, O., Gadawska, I., Cote, H.C.F., and Hsieh, A.Y.Y. (2020). Platelet mtDNA content and leukocyte count influence whole blood mtDNA content. Mitochondrion 52, 108–114.

Spinelli, J.B., and Haigis, M.C. (2018). The multifaceted contributions of mitochondria to cellular metabolism. Nat Cell Biol 20, 745–754.

Stewart, J.B., and Chinnery, P.F. (2021). Extreme heterogeneity of human mitochondrial DNA from organelles to populations. Nat Rev Genet 22, 106–118.

Sun, N., Youle, R.J., and Finkel, T. (2016). The Mitochondrial Basis of Aging. Mol Cell 61, 654–666.

Thul, P.J., Akesson, L., Wiking, M., Mahdessian, D., Geladaki, A., Ait Blal, H., Alm, T., Asplund, A., Bjork, L., Breckels, L.M., et al. (2017). A subcellular map of the human proteome. Science 356.

Vaillant-Beuchot, L., Mary, A., Pardossi-Piquard, R., Bourgeois, A., Lauritzen, I., Eysert, F., Kinoshita, P.F., Cazareth, J., Badot, C., Fragaki, K., et al. (2021). Accumulation of amyloid precursor protein C-terminal fragments triggers mitochondrial structure, function, and mitophagy defects in Alzheimer’s disease models and human brains. Acta Neuropathol 141, 39–65.

Wachsmuth, M., Hubner, A., Li, M., Madea, B., and Stoneking, M. (2016). Age-Related and Heteroplasmy-Related Variation in Human mtDNA Copy Number. PLoS Genet 12, e1005939.

Wang, M., Beckmann, N.D., Roussos, P., Wang, E., Zhou, X., Wang, Q., Ming, C., Neff, R., Ma, W., Fullard, J.F., et al. (2018). The Mount Sinai cohort of large-scale genomic, transcriptomic and proteomic data in Alzheimer’s disease. Sci Data 5, 180185.

Wang, W., Li, L., Lin, W.L., Dickson, D.W., Petrucelli, L., Zhang, T., and Wang, X. (2013). The ALS disease-associated mutant TDP-43 impairs mitochondrial dynamics and function in motor neurons. Hum Mol Genet 22, 4706–4719.

Wang, W., Wang, L., Lu, J., Siedlak, S.L., Fujioka, H., Liang, J., Jiang, S., Ma, X., Jiang, Z., da Rocha, E.L., et al. (2016). The inhibition of TDP-43 mitochondrial localization blocks its neuronal toxicity. Nat Med 22, 869–878.

Wang, X., Allen, M., Li, S., Quicksall, Z.S., Patel, T.A., Carnwath, T.P., Reddy, J.S., Carrasquillo, M.M., Lincoln, S.J., Nguyen, T.T., et al. (2020). Deciphering cellular transcriptional alterations in Alzheimer’s disease brains. Mol Neurodegener 15, 38.

Wei, W., Keogh, M.J., Wilson, I., Coxhead, J., Ryan, S., Rollinson, S., Griffin, H., Kurzawa-Akanbi, M., Santibanez-Koref, M., Talbot, K., et al. (2017). Mitochondrial DNA point mutations and relative copy number in 1363 disease and control human brains. Acta Neuropathol Commun 5, 13.

Wingo, A.P., Fan, W., Duong, D.M., Gerasimov, E.S., Dammer, E.B., Liu, Y., Harerimana, N.V., White, B., Thambisetty, M., Troncoso, J.C., et al. (2020). Shared proteomic effects of cerebral atherosclerosis and Alzheimer’s disease on the human brain. Nat Neurosci 23, 696–700.

Yang, S.Y., Castellani, C.A., Longchamps, R.J., Pillalamarri, V.K., O’Rourke, B., Guallar, E., and Arking, D.E. (2021). Blood-derived mitochondrial DNA copy number is associated with gene expression across multiple tissues and is predictive for incident neurodegenerative disease. Genome Res 31, 349–358.

Yin, J., Reiman, E.M., Beach, T.G., Serrano, G.E., Sabbagh, M.N., Nielsen, M., Caselli, R.J., and Shi, J. (2020). Effect of ApoE isoforms on mitochondria in Alzheimer disease. Neurology 94, e2404–e2411.

Yu, C.H., Davidson, S., Harapas, C.R., Hilton, J.B., Mlodzianoski, M.J., Laohamonthonkul, P., Louis, C., Low, R.R.J., Moecking, J., De Nardo, D., et al. (2020). TDP-43 Triggers Mitochondrial DNA Release via mPTP to Activate cGAS/STING in ALS. Cell 183, 636–649 e618.

Zhang, R., Wang, Y., Ye, K., Picard, M., and Gu, Z. (2017). Independent impacts of aging on mitochondrial DNA quantity and quality in humans. BMC Genomics 18, 890.

Zhong, Y., Wan, Y.W., Pang, K., Chow, L.M., and Liu, Z. (2013). Digital sorting of complex tissues for cell type-specific gene expression profiles. BMC Bioinformatics 14, 89.

