## Supplemental Tables and Figures for "Characterization of mitochondrial DNA quantity and quality in the human aged and Alzheimer’s disease brain"

**Table S1. Characteristics of the Mayo cohort.**

| | Controls (n=51) | AD (n=89) | PSP (n=83) | Pathologic aging (n=39) |
|---|---|---|---|---|
| Sex (male) | 24 (47.1%) | 34 (38.2%) | 50 (60.2%) | 15 (38.5%) |
| Age (years) | 88.4 (12.1) | 84.3 (9.9) | 74.1 (6.4) | 90.3 (9.4) |
| Post mortem interval (hours) | 10.3 (8.7) | 7.1 (5.8) | 8.7 (6.1) | - |

Mean and standard deviation are shown for the continuous variables age and post mortem interval. Since age was right-censored at 95 years, a Tobit model was used to estimate mean and standard deviation of age.

**Table S2. Characteristics of the MSBB cohort.**

| | Controls (n=132) | AD (n=205) |
|---|---|---|
| Sex (male) | 47 (35.6%) | 71 (34.6%) |
| Age (years) | 84.7 (11.4) | 86.5 (10.7) |
| Post mortem interval (hours) | 9.1 (6.6) | 6.5 (4.6) |

Mean and standard deviation are shown for the continuous variables age and post mortem interval. Since age was right-censored at 95 years, a Tobit model was used to estimate mean and standard deviation of age.

**Table S3. Detailed results from the univariate regression analyses shown in Fig. 2A.**

| | DLPFC | | | | | PCC | | | | | CB | | | | |
|---|---|---|---|---|---|---|---|---|---|---|---|---|---|---|---|
| | n | $R^2_{part}$ | $p_F$ | β | $p_t$ | n | $R^2_{part}$ | $p_F$ | β | $p_t$ | n | $R^2_{part}$ | $p_F$ | β | $p_t$ |
| Pathologic AD | 454 | 0.030 | 0.00023 | | | 66 | 0.043 | 0.098 | | | 242 | 0.004 | 0.36 | | |
| *No AD* | 146 | | | | | 21 | | | | | 101 | - | - | | |
| *AD* | 308 | | | -0.37 | 0.00023 | 45 | | | -0.45 | 0.098 | 141 | - | - | -0.13 | 0.36 |
| Global AD path. | 454 | 0.056 | $3.3\times10^{-7}$ | -0.24 | $3.3\times10^{-7}$ | 66 | 0.015 | 0.34 | -0.12 | 0.34 | 242 | 0.015 | 0.059 | -0.13 | 0.059 |
| Aymloid | 451 | 0.015 | 0.011 | -0.12 | 0.011 | 66 | 0.003 | 0.66 | -0.06 | 0.66 | 242 | 0.011 | 0.1 | -0.11 | 0.10 |
| Tau | 450 | 0.048 | $2.9\times10^{-6}$ | -0.22 | $2.9\times10^{-6}$ | 66 | 0.033 | 0.15 | -0.19 | 0.15 | 242 | 0.005 | 0.28 | -0.07 | 0.28 |
| TDP-43 | 422 | 0.011 | 0.21 | | | 63 | 0.226 | 0.0021 | | | 212 | 0.014 | 0.41 | | |
| *None* | 205 | | | | | 23 | | | | | 96 | - | - | | |
| *Amygdala* | 76 | | | -0.05 | 0.70 | 16 | | | -0.43 | 0.17 | 45 | - | - | 0.00 | 0.98 |
| *Limbic* | 97 | | | -0.02 | 0.90 | 12 | | | -0.59 | 0.092 | 38 | - | - | -0.01 | 0.95 |
| *Neocortical* | 44 | | | -0.35 | 0.039 | 12 | | | -1.32 | 0.00016 | 33 | - | - | 0.30 | 0.12 |
| Lewy bodies | 436 | 0.008 | 0.34 | | | 61 | 0.028 | 0.66 | | | 236 | 0.003 | 0.87 | | |
| *None* | 337 | | | | | 48 | | | | | 184 | - | - | | |
| *Nigral* | 7 | | | 0.19 | 0.62 | 2 | | | 0.91 | 0.22 | 6 | - | - | -0.22 | 0.60 |
| *Limbic* | 32 | | | -0.12 | 0.53 | 2 | | | 0.23 | 0.76 | 21 | - | - | 0.08 | 0.74 |
| *Neocortical* | 60 | | | -0.23 | 0.098 | 9 | | | 0.06 | 0.87 | 25 | - | - | -0.12 | 0.59 |
| CAA | 445 | 0.035 | 0.0013 | | | 65 | 0.132 | 0.038 | | | 234 | 0.037 | 0.034 | | |
| *None* | 92 | | | | | 13 | | | | | 58 | - | - | | |
| *Mild* | 191 | | | -0.03 | 0.81 | 25 | | | -0.29 | 0.39 | 101 | - | - | 0.27 | 0.10 |
| *Moderate* | 110 | | | -0.17 | 0.23 | 16 | | | -0.69 | 0.066 | 51 | - | - | 0.48 | 0.014 |
| *Severe* | 52 | | | -0.61 | 0.00042 | 11 | | | -1.09 | 0.0086 | 24 | - | - | -0.09 | 0.72 |
| Cereb. Atheroscl. | 452 | 0.005 | 0.49 | | | 66 | 0.007 | 0.94 | | | 240 | 0.033 | 0.048 | | |
| *None* | 84 | | | | | 11 | | | | | 39 | - | - | | |
| *Mild* | 212 | | | -0.03 | 0.84 | 25 | | | 0.13 | 0.73 | 114 | - | - | 0.35 | 0.06 |
| *Moderate* | 123 | | | 0.05 | 0.74 | 19 | | | -0.06 | 0.88 | 72 | - | - | 0.48 | 0.017 |
| *Severe* | 33 | | | -0.25 | 0.22 | 11 | | | -0.03 | 0.95 | 15 | - | - | -0.06 | 0.85 |
| Arteriolosclerosis | 453 | 0.011 | 0.18 | | | 66 | 0.039 | 0.49 | | | 239 | 0.008 | 0.62 | | |
| *None* | 136 | | | | | 9 | | | | | 78 | - | - | | |
| *Mild* | 157 | | | -0.06 | 0.61 | 26 | | | -0.10 | 0.80 | 79 | - | - | -0.04 | 0.83 |
| *Moderate* | 118 | | | -0.02 | 0.87 | 22 | | | -0.50 | 0.22 | 65 | - | - | 0.12 | 0.47 |
| *Severe* | 42 | | | -0.38 | 0.033 | 9 | | | -0.31 | 0.52 | 17 | - | - | 0.25 | 0.35 |
| Gross infarcts | 454 | 0.008 | 0.059 | | | 66 | 0.008 | 0.48 | | | 242 | 0.001 | 0.71 | | |
| *None* | 283 | | | | | 44 | | | | | 159 | - | - | | |
| *One or more* | 171 | | | -0.18 | 0.059 | 22 | | | -0.19 | 0.48 | 83 | - | - | 0.05 | 0.71 |
| Microinfarcts | 454 | 0.007 | 0.071 | | | 66 | 0.128 | 0.0037 | | | 242 | 0.005 | 0.27 | | |
| *None* | 322 | | | | | 48 | | | | | 172 | - | - | | |
| *One or more* | 132 | | | -0.19 | 0.071 | 18 | | | -0.80 | 0.0037 | 70 | - | - | 0.16 | 0.27 |
| Hippocam. scler. | 449 | 0.001 | 0.64 | | | 65 | 0.027 | 0.20 | | | 240 | 0.014 | 0.069 | | |
| *Not present* | 410 | | | | | 59 | | | | | 218 | - | - | | |
| *present* | 39 | | | -0.08 | 0.64 | 6 | | | -0.57 | 0.2 | 22 | - | - | 0.41 | 0.069 |
| Global cognition | 453 | 0.081 | $7.9\times10^{-10}$ | 0.29 | $7.9\times10^{-10}$ | 66 | 0.264 | $1.7\times10^{-5}$ | 0.56 | $1.7\times10^{-5}$ | 239 | 0.003 | 0.42 | 0.05 | 0.42 |
| Cognitive decline | 437 | 0.080 | $1.8\times10^{-9}$ | 0.29 | $1.8\times10^{-9}$ | 62 | 0.191 | 0.00055 | 0.45 | 0.00055 | 208 | 0.001 | 0.65 | 0.03 | 0.65 |
| Age | 454 | 0.002 | 0.32 | 0.05 | 0.32 | 66 | 0.004 | 0.61 | -0.06 | 0.61 | 242 | 0.005 | 0.27 | -0.07 | 0.27 |
| Sex | 454 | 0.001 | 0.60 | | | 66 | 0.017 | 0.30 | | | 242 | 0.001 | 0.58 | | |
| *Female* | 296 | | | | | 45 | | | | | 171 | - | - | | |
| *Male* | 158 | | | 0.05 | 0.60 | 21 | | | -0.28 | 0.30 | 71 | - | - | -0.08 | 0.58 |

Each pathologic variable was analyzed separately in a regression model adjusted only for age and sex. Global cognition and cognitive decline were analyzed separately in a regression model adjusted only for age, sex, and education. Age and sex were analyzed in a regression model adjusted for global AD pathology.

**Table S4. Association between cell type proportions and mtDNAcn in the DLPFC (n=327).**

| | $R^2_{part}$ | β | p |
|---|---|---|---|
| Neurons | 0.069 | 0.23 | $1.7 \times 10^{-6}$ |
| Astrocytes | 0.002 | -0.37 | 0.48 |
| Oligodendrocytes | 0.019 | -0.12 | 0.013 |
| Microglia | 0.001 | -0.02 | 0.66 |
| Endothelial cells | 0.003 | -0.05 | 0.30 |

Partial $R^2$, coefficient for the cell type, and p value were derived from separate linear regression models with mtDNAcn as outcome controlled for age and sex.

**Table S5. Association between number of mtDNA heteroplasmic mutations and age adjusted for pathologic diagnosis.**

| | DLPFC/ROSMAP (n=454) | | | TCX/Mayo (n=262) | | | FP/MSBB (n=270) | | |
|---|---|---|---|---|---|---|---|---|---|
| | β | SE | p | β | SE | p | β | SE | p |
| Age | 0.015 | 0.004 | $1.4\times10^{-5}$ | 0.017 | 0.004 | $2.1\times10^{-5}$ | 0.016 | 0.004 | $4.4\times10^{-5}$ |
| Sex | 0.022 | 0.048 | 0.64 | 0.058 | 0.058 | 0.32 | 0.035 | 0.067 | 0.60 |
| AD | 0.019 | 0.049 | 0.70 | 0.184 | 0.082 | 0.026 | 0.125 | 0.063 | 0.051 |
| Path. aging | | | | 0.079 | 0.097 | 0.42 | | | |
| PSP | | | | 0.136 | 0.098 | 0.17 | | | |
| mtDNAcn | 0.037 | 0.023 | 0.11 | 0.100 | 0.031 | 0.0013 | 0.014 | 0.031 | 0.66 |

PSP = Progressive supranuclear palsy, Path. aging = Pathologic aging

**Table S6. Datasets and analysis results deposited at the AD Knowledge Portal.**

| Study | Data description | File type | Accession number |
|---|---|---|---|
| ROSMAP | Whole-genome sequencing data | bam | syn20068543 |
| | Nuclear variants | vcf | syn11724057 |
| | Mt variants | vcf | syn25705298 |
| | mtDNAcn and heteroplasmy levels | csv | syn25705303 |
| | TMT proteomic data | csv | syn21266454 |
| | RNA-seq data | bam | syn21188662 |
| Mayo | Whole-genome sequencing data | bam | syn19989379 |
| | Nuclear variants | vcf | syn11724002 |
| | Mt variants | vcf | syn25705298 |
| | mtDNAcn and heteroplasmy levels | csv | syn25705303 |
| MSBB | Whole-genome sequencing data | bam | syn19987071 |
| | Nuclear variants | vcf | syn11723899 |
| | Mt variants | vcf | syn25705298 |
| | mtDNAcn and heteroplasmy levels | csv | syn25705303 |

All datasets can be accessed via the manuscript's website at the AD knowledge portal: https://doi.org/10.7303/syn25618990

### Supplementary Figures

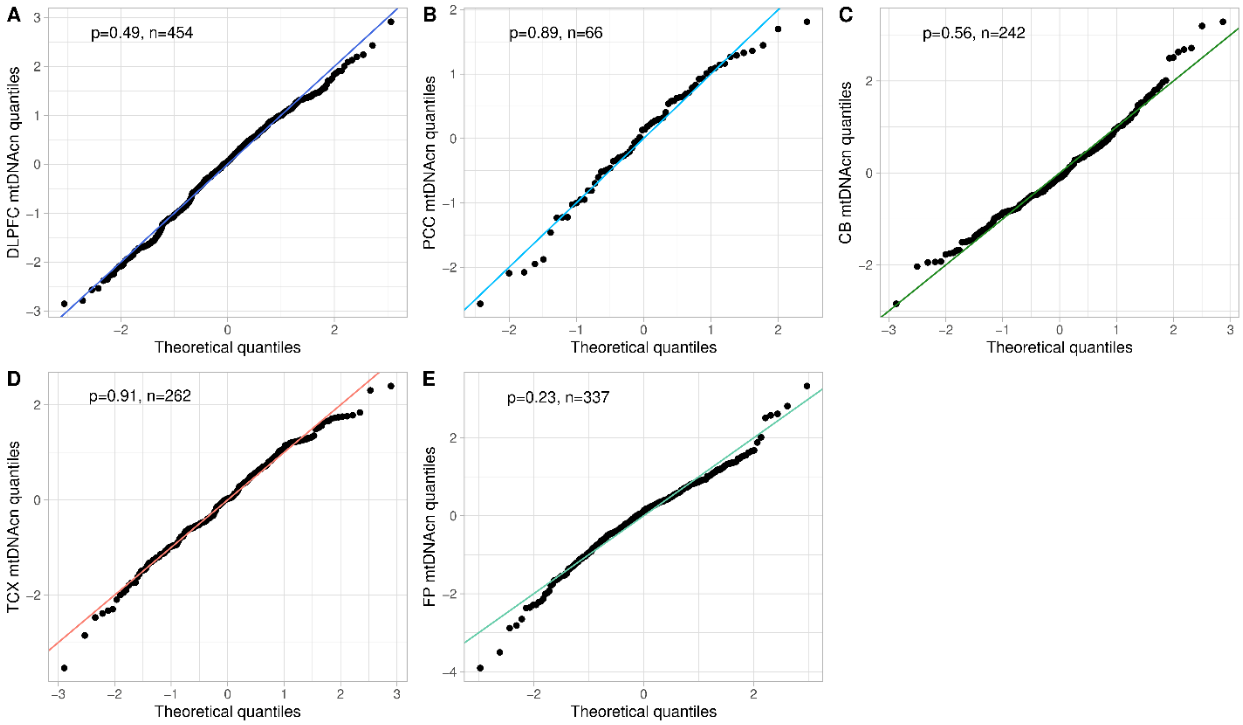


**Figure S1. Log-transformed mtDNAcn values are approximately normally distributed.**
(A-E) Plots show the observed quantiles of the log-transformed and standardized mtDNAcn values versus the quantiles of the standard normal distribution for the DLPFC samples from ROSMAP (A), the PCC samples from ROSMAP (B), the CB samples from ROSMAP (C), the TCX samples from Mayo (D), and the FP samples from MSBB (E). P values were calculated using the Kolmogorov-Smirnov test.

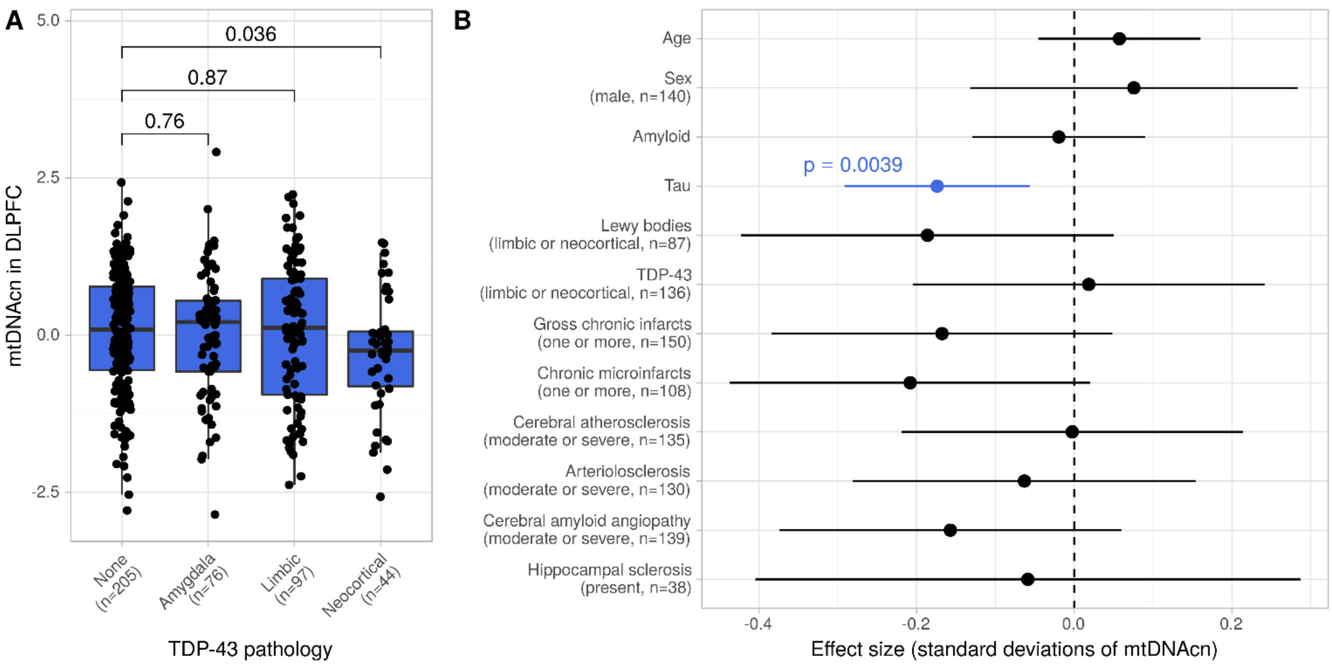


**Figure S2: Changes of the mtDNAcn are primarily associated with tau in the DLPFC.**

(B) Boxplot shows the mtDNAcn in the DLPFC for different stages of TDP-43 pathology. Wilcoxon rank-sum test was used to calculate p values.

(D) Forest plot shows the result from a multivariable regression model with mtDNAcn as outcome and the pathologic and demographic variables denoted on the y axis as explanatory variables. Estimated coefficients are shown as dots and the line segments represent the respective 95% confidence intervals. Continuous variables were z-standardized. Categorical variables were dichotomized and the factor level and case numbers corresponding to the plotted coefficient are denoted in brackets under the variable name. t-test was applied to calculate p values. A total of n=394 cases with complete observations of all variables were used to fit the model.

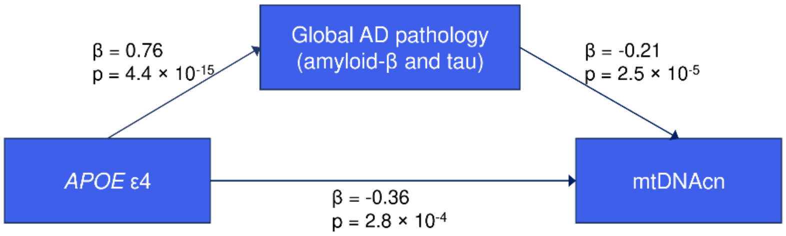


**Figure S3: Significant fraction of the *APOE* ε4 effect on mtDNAcn is mediated via AD pathologies.**
Mediation analysis was performed assuming global AD pathology (quantitative summary of neuritic plaques, diffuse plaques, and neurofibrillary tangles) mediates parts of the total effect of *APOE* ε4 on mtDNAcn. The total effect is represented by the lower arrow in the figure. In this model, the upper path via global AD pathology (average causal mediation effects) was significant ($\beta=-0.16$, $p\leq10^{-16}$) and represented 44% of the total effect. The average direct effects were estimated to be $\beta=-0.20$ and achieved borderline significance ($p=0.051$) (see Table 2 in the main manuscript for details). The analysis conducted on the 454 samples from the DLPFC. Global AD pathology and mtDNAcn were standardized for better interpretability of the effect size. *APOE* ε4 effect sizes are in dosage of the ε4 allele.

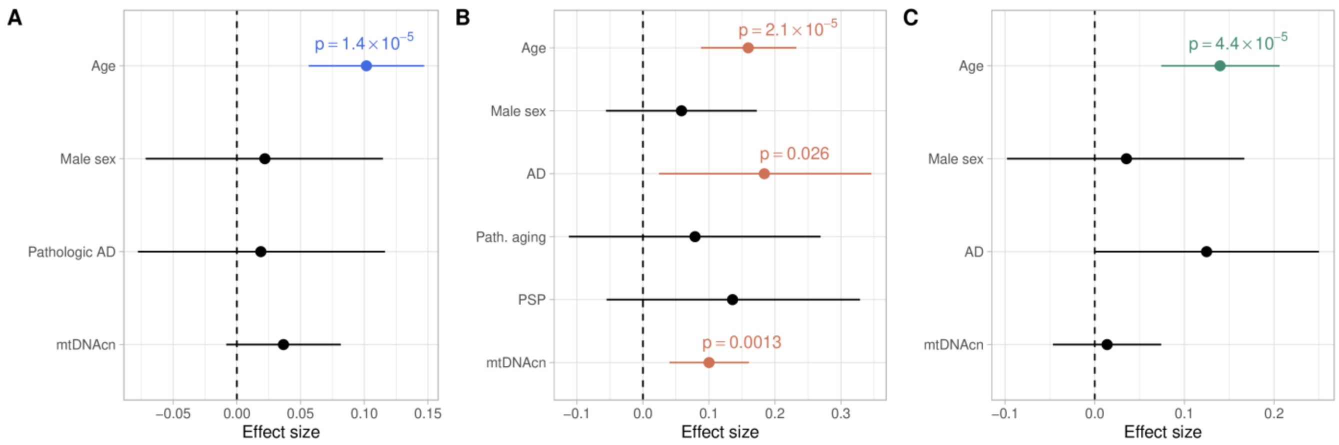


**Figure S4. Number of mtDNA heteroplasmic mutations in cortical regions is associated with age adjusted for sex, pathologic diagnosis, and mtDNAcn.**

(A) Forest plot shows results from a quasi-Poisson regression model with the number of mtDNA heteroplasmic mutations in the DLPFC (n=454) as outcome. For better comparability of the effect sizes, age and mtDNAcn were z-standardized before fitting the model (i.e., the effect size shown for age is the change in standard deviations of mtDNAcn per standard deviation of age). P values of significant variables are indicated in blue.

(B) Similar plot as (A) showing results from the TCX data (n=262) from the Mayo study. In addition to controls and AD samples, the Mayo study included samples with pathologic aging and progressive supranuclear palsy (PSP). P values of significant variables are indicated in orange.

(C) Similar plot as (A) showing results from the FP data (n=270) from the MSBB study. P values of significant variables are indicated in green.

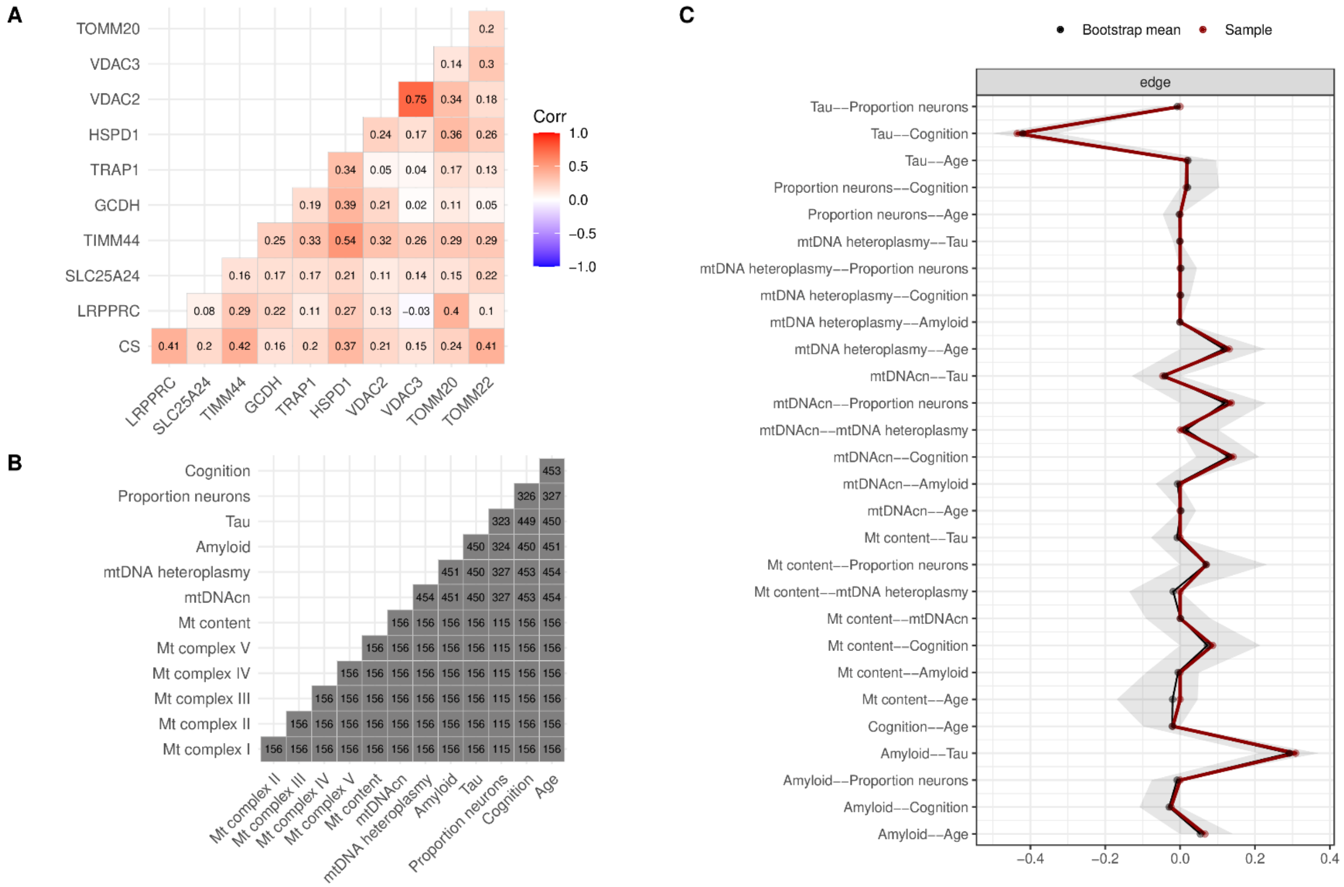


**Figure S5. Mitochondrial content score and its relation to mtDNAcn, mtDNA heteroplasmy burden and AD-related phenotypes.**

(A) Heatmap shows pairwise Pearson correlations between the 10 selected proteins that were used to calculate the mitochondrial content score in the DLPFC (n=156).

(B) The lower triangular matrix matches Fig. 5A in the main manuscript and depicts the number of samples with pairwise complete observations that were used to calculate the Pearson correlations shown in Fig. 5A.

(C) The figure visualizes bootstrapping results to assess the stability of the partial correlations shown in the graph in Fig. 5B in the main manuscript. All possible pairs of variables (edges in the graph) are shown on the y axis. The x axis shows the partial correlation of the full sample (red), the mean over all 1,000 bootstraps (dark gray), and the 95% confidence interval derived from the bootstraps (light gray band).
